# Exercise Training Improves Skeletal Muscle Insulin Sensitivity and Reprograms the Adipose Transcriptome in Heavier Monozygotic Twins

**DOI:** 10.64898/2026.06.15.26355744

**Authors:** Jaakko Hentilä, Max Ullrich, Ronja Ojala, Martin S. Lietzén, Marja A. Heiskanen, Thibaux Van der Stede, Sanna Honkala, Mika Helmiö, Johan Rajander, Olli Eskola, Eliisa Löyttyniemi, Riikka Lautamäki, Heidi Virtanen, Kalle Koskensalo, Olli J. Heinonen, Kirsi H. Pietiläinen, Jaakko Kaprio, Riikka Kivelä, Adam P. Sharples, Jarna C. Hannukainen

## Abstract

Exercise training improves skeletal muscle insulin sensitivity, yet its effects on white adipose tissue remain incompletely understood. We investigated how adiposity and exercise training influence insulin-stimulated glucose uptake in skeletal muscle and abdominal subcutaneous adipose tissue (ASAT), alongside adaptations in gene expression and DNA-methylation. Ten monozygotic twin pairs discordant for BMI underwent [^18^F]FDG-PET/CT imaging of skeletal muscle (vastus lateralis, VL) and ASAT during a euglycemic-hyperinsulinaemic clamp before and after six months of exercise training. VL and ASAT biopsies were analyzed using mRNA-sequencing and reduced representation bisulfite sequencing.

Exercise training improved whole-body and VL insulin sensitivity in leaner and heavier co-twins (p<0.05), without altering ASAT insulin sensitivity or body weight. Whole body adiposity exerted a stronger impact on ASAT molecular profiles than on skeletal muscle. At baseline, heavier co-twins displayed widespread ASAT transcriptional alterations enriched for inflammatory, proliferative and extracellular matrix pathways compared with leaner co-twins. In heavier co-twins, exercise training attenuated inflammatory and proliferative signatures in ASAT and induced transcriptomic convergence with the leaner co-twins. These changes were accompanied by marked shifts in transcription factor activity and context-specific DNA methylation changes. In contrast, VL exhibited more modest transcriptomic and epigenetic responses relative to ASAT, particularly in heavier co-twins.

In conclusion, six months of exercise training improved whole-body and VL insulin sensitivity while in ASAT many of the obesity associated transcriptomic programmes were reversed. These findings highlight adipose tissue as a major site of obesity– and exercise-responsive molecular plasticity and reveal tissue-specific regulatory mechanisms that contribute to the metabolic benefits of exercise training.

*Clinical Trial Registration Number: NCT03730610.* https://clinicaltrials.gov/study/NCT03730610

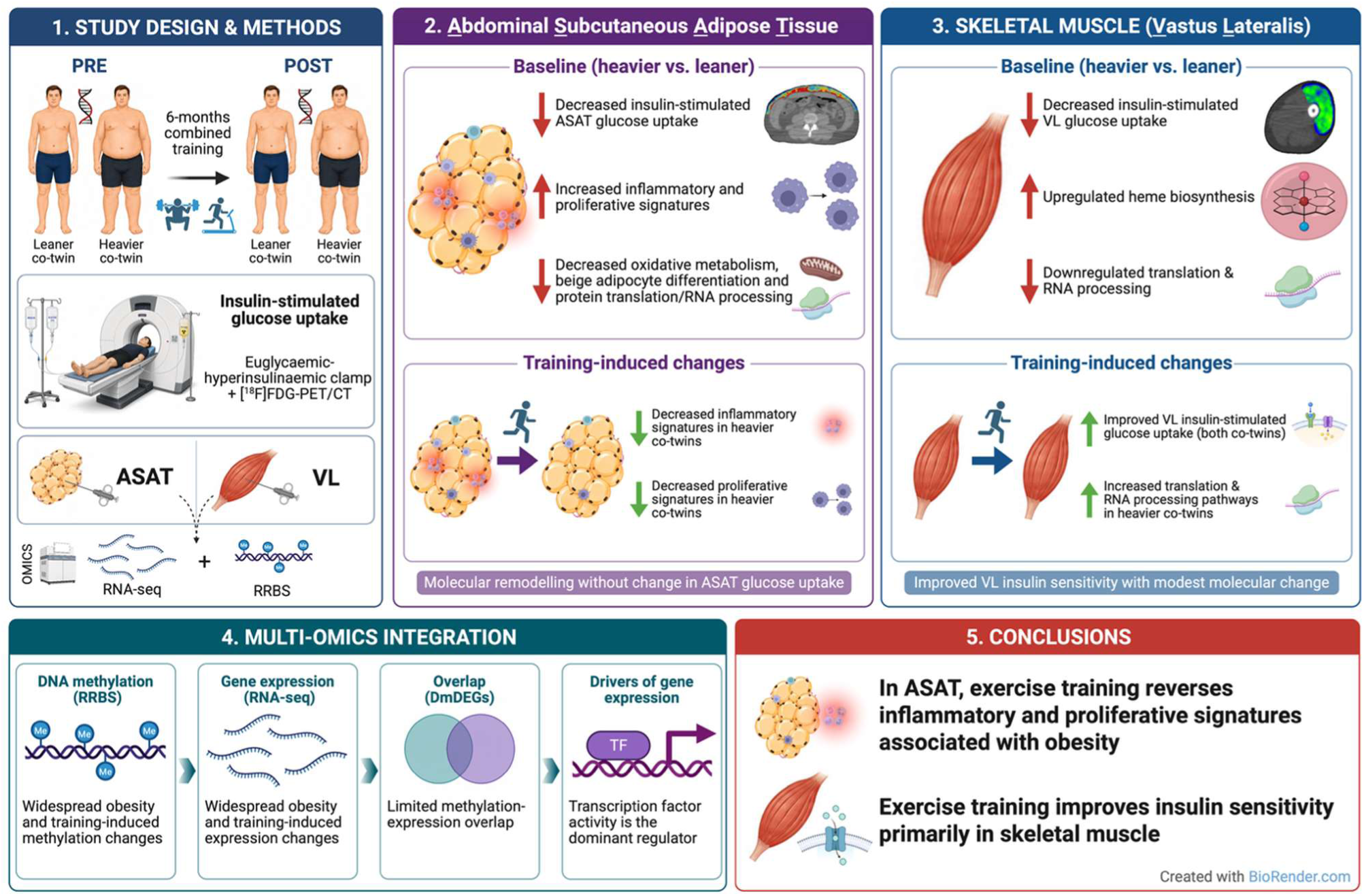

## 1 INTRODUCTION

Obesity is associated with metabolic disorders such as type 2 diabetes (T2D), which often begins with peripheral insulin resistance, an attenuated physiological response to insulin[1,2]. A key contributor to insulin resistance is low-grade inflammation originating from expanding adipose tissue during energy surplus, although the underlying mechanisms remain unclear[3,4]. Proposed triggers include hypoxia, adipocyte apoptosis, and mechanical stress, which may initiate an inflammatory response during rapid adipose tissue expansion[3]. This inflammation can induce local insulin resistance, initially protecting adipocytes from pathologic growth[3], yet at the cost of impaired triglyceride storage and increased release of free fatty acids into the bloodstream. Consequently, ectopic fat accumulates in and around internal organs and peripheral tissues such as skeletal muscle, where it exerts lipotoxic effects that further promote insulin resistance, a major risk factor for metabolic diseases[5].

Previous omics-based approaches have identified several molecular pathways in skeletal muscle and adipose tissue that are altered with changes in adiposity, providing insights into the mechanisms underlying obesity-related tissue impairments. For example, transcriptomic studies in skeletal muscle from individuals with obesity or T2D reveal downregulation of genes involved in mitochondria, insulin signaling, and amino acid metabolism alongside upregulation of genes related to N-glycan biosynthesis, apoptosis and inflammation[6,7]. Furthermore, lifestyle-induced weight gain has been associated with upregulation of genes involved in inflammation, cell differentiation and extracellular matrix (ECM) remodeling, whereas downregulated genes have been primarily related to mitochondrial and RNA metabolism[6,8]; notably obesity elicits more pronounced gene expression changes in adipose tissue than in skeletal muscle[6].

One of the mechanisms regulating gene expression at the transcriptional level is DNA methylation at gene regulatory regions such as promoters, which often contain cytosine-phosphate-guanine (CpG) rich regions. In this process, methyl groups are added or removed on cytosine residues within CpG dinucleotides typically leading to decreased and increased gene expression respectively[9]. Current evidence suggests that obesity-associated changes in gene expression in adipose tissue and skeletal muscle may be related to DNA methylation[10,11], although the mechanistic links have not yet been fully defined. Also, the impact of adiposity-driven alterations in gene expression on tissue-specific and systemic insulin-sensitivity remains incompletely understood.

Overwhelming evidence indicates that exercise training is beneficial in the prevention and treatment of insulin resistance and T2D, as it improves whole-body insulin sensitivity primarily through adaptations in skeletal muscle tissue[12–14]. The exercise-induced improvement in skeletal muscle insulin sensitivity is multifactorial, and some mechanisms remain incompletely understood. Current evidence suggests that exercise training enhances mitochondrial function, reduces lipotoxic derivatives and alters the cytoskeletal architecture of skeletal muscle[15–17], which may facilitate glucose transport from the blood into muscle tissue[1]. Preclinical and human studies indicate that exercise training may improve whole-body insulin sensitivity through adipose tissue remodeling, including reductions in inflammation and increases in vascularization[18–20]. Furthermore, studies have shown that exercise training induces epigenetic modifications in skeletal muscle[21–27] and adipose tissue[28,29]. These modifications may regulate the expression of genes involved in tissue remodeling, although their precise role in tissue-specific and whole-body glucose homeostasis remains unknown.

Previous studies using [^18^F]FDG-positron emission tomography/computed tomography (PET/CT) imaging during euglycaemic-hyperinsulinaemic clamp have shown that body adiposity is associated with lower insulin-stimulated glucose uptake in white adipose[20] and skeletal muscle tissue[30]. Furthermore, exercise training has been shown to enhance insulin-stimulated glucose uptake in skeletal muscle among individuals who are moderately overweight[31], prediabetic or have T2D[32]. However, the effect of exercise training on insulin-stimulated glucose uptake in white adipose tissue remains unclear, as two studies reported depot-specific improvements[20,33], whereas one found no significant change[31]. Although glucose uptake in adipose tissue and skeletal muscle has been examined multiple times in individuals with obesity and following exercise training by [^18^F]FDG-PET[14,20,30,32,33], few studies have concurrently investigated the molecular adaptations underlying these changes. Additionally, the studies that have analyzed molecular adaptations have focused predominantly on pre-determined genes, molecular pathways or biomarkers[20,31].

To address this gap, monozygotic twin pairs discordant for BMI were studied to control for genetic factors, allowing us to examine the effect of increased body weight on insulin-stimulated glucose uptake in both skeletal muscle and abdominal subcutaneous adipose tissue (ASAT) using [^18^F]FDG-PET. Importantly, we investigated whether six months of a combined multi-modal exercise training intervention incorporating aerobic, resistance, and high-intensity interval training could reverse the obesity-associated impairments in insulin-stimulated glucose uptake in skeletal muscle and ASAT. To obtain a comprehensive view of obesity– and exercise training-induced molecular adaptations underlying insulin-stimulated glucose uptake, we analyzed the transcriptome (RNA-seq), DNA methylome (RRBS) and inferred transcription factor (TF) activity from RNA-Seq data in skeletal muscle and ASAT before and after the 6-month training intervention.

## 2 MATERIALS AND METHODS

### 2.1 Ethics

This study was conducted as part of a clinical exercise training intervention entitled: “Systemic cross-talk between brain, gut and peripheral tissues in glucose homeostasis: effects of exercise training” (CROSSYS, ClinicalTrials.gov identifier NCT03730610). The study protocol, participant information and informed consent procedures were approved by the Ethical Committee of the Hospital District of Southern Finland (approval numbers 100/1801/2018/438§ and 548§). Participants had the right to refuse any part of the study protocol or discontinue at any time without the need to provide any explanation. All procedures adhered to Good Clinical Practice guidelines, were performed within Finnish law and the principles outlined in the Declaration of Helsinki.

### 2.2 Participants and design

Monozygotic twin pairs discordant for BMI were recruited from three population-based longitudinal twin cohort studies[34]. The inclusion criteria were a BMI within-pair difference of ≥ 2 kg/m^2^ and at least one of the co-twins had to be overweight (BMI > 25 kg/m^2^). The exclusion criteria are previously detailed in our protocol paper[34]. Of the 12 pairs enrolled (age mean 40.4 years, 95% CI [37.5; 43.4]; leaner co-twins BMI: mean 29.1 kg/m^2^, 95% CI [25.2; 33.0], heavier co-twins BMI: mean 36.7 kg/m^2^,

95% CI [32.7; 40.7]; 8 female pairs) 10 completed the training intervention (Figure 1A). Based on American Diabetes Association criteria, both co-twin groups were classified as non-diabetic; however, five leaner co-twins and seven heavier co-twins had impaired fasting glucose, and two individuals in each group had impaired glucose tolerance. The smoking status was the same in 9 pairs and differed in 2 pairs (in both pairs the leaner co-twin smoked). In one pair the heavier co-twin smoked and the leaner co-twin used snus. There were no differences in alcohol consumption between co-twins at baseline. During the screening visit, participants underwent anthropometric measurements, a health examination and an oral glucose tolerance test (OGTT) following an overnight fast (10 h), along with exercise performance tests at Paavo Nurmi Centre (Turku, Finland). Baseline assessments were conducted a few weeks later over two consecutive days (Figure 1B), including [^18^F]FDG-PET/CT during euglycaemic-hyperinsulinaemic clamp, whole-body MRI and biopsies of skeletal muscle and subcutaneous adipose tissue (detailed below). Participants then completed a six month home-based exercise intervention (detailed below), after which all baseline measurements were repeated (Figure 1B)[34].

**Figure 1.**
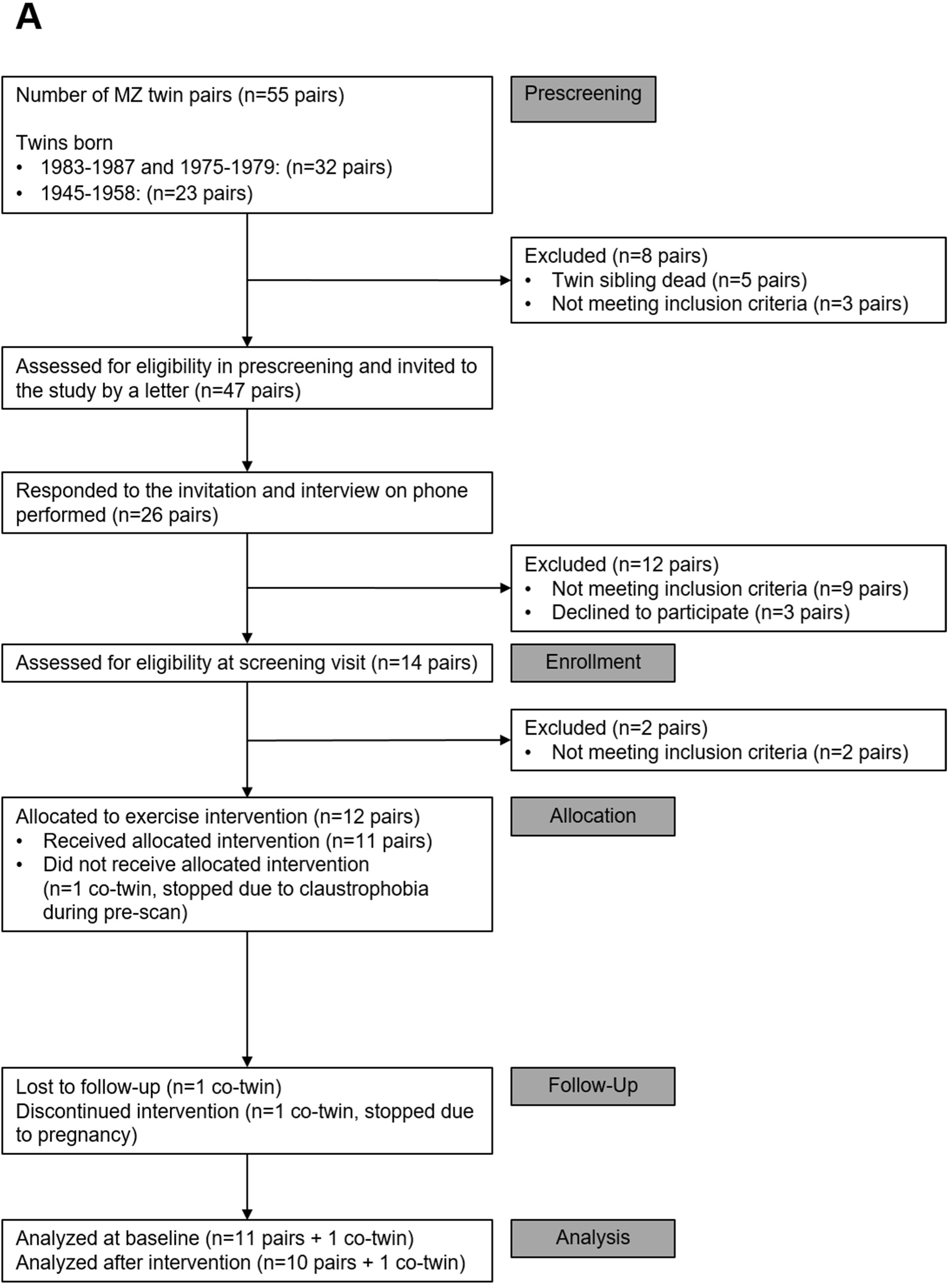
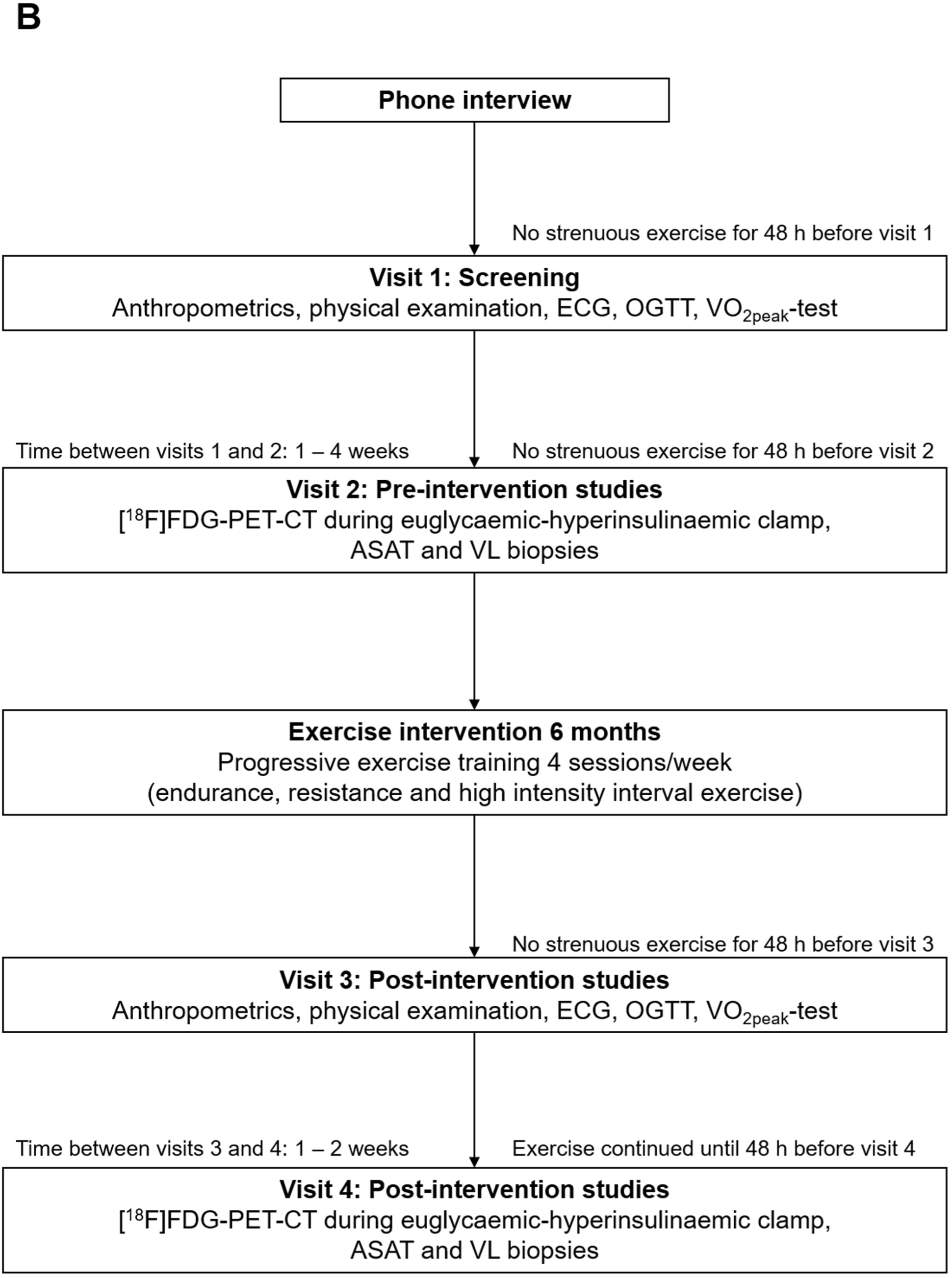
A) Consort flow chart of the study. MZ=monozygotic. B) Overview of the study protocol. ECG: electrocardiography, OGTT: oral glucose tolerance test, VO2peak: peak oxygen uptake, [^18^F]FDG: 2-deoxy-2-[^18^F]fluoro-D-glucose, PET: positron emission tomography, CT: computed tomography, MRI: magnetic resonance imaging, ASAT: abdominal subcutaneous adipose tissue, VL: vastus lateralis

### 2.3 Exercise performance tests, anthropometric measurements, and training intervention

Cardiorespiratory fitness (VO_2peak_) was assessed using a stationary bicycle ergometer test (Ergoline 800 s, VIASYS Healthcare Germany) until exhaustion and body composition was measured using Inbody 720 (Biospace Co, Korea) at the Paavo Nurmi Centre (Turku, Finland) as previously described[34]. The six-month exercise training involved progressive training, including two endurance sessions, one resistance session, and one high-intensity exercise (HIIE) session as described in our protocol[34]. Participants trained at home due to their dispersed locations across Finland, with one weekly session, typically resistance or high-intensity exercise (HIIE), supervised by a qualified personal trainer. Training intensity, adherence, and progression were monitored using heart rate monitors (Polar A370, Polar, Finland) and training logs[34]. For endurance training, both intensity (%HRmax) and volume (duration per exercise) increased progressively as previously described[34]. For resistance training, the repetitions and sets remained constant, while the load was progressively increased and exercises were advanced to more challenging variations. Further methodological details are provided in our published protocol[34]. Participants completed a food diary for three consecutive days before (PRE) and after (POST) the intervention and dietary intake was analyzed using Fineli software (https://fineli.fi/fineli/fi/index) to determine total energy intake. Visceral fat mass was analyzed as previously described[35].

### 2.4 Euglycaemic-hyperinsulinaemic clamp and [^18^F]FDG-PET/CT scan

Insulin-stimulated glucose uptake was assessed during a euglycaemic-hyperinsulinaemic clamp in VL skeletal muscle and ASAT using [^18^F]FDG-PET/CT (Discovery MI, GE Healthcare, US)[34]. The M-value (whole-body glucose disposal rate) was calculated as previously described[35]. Once steady state was achieved during the clamp, [^18^F]FDG (mean 155.47, SD 7.76 MBq) was injected to the antecubital vein via a catheter. PET/CT scans of the abdominal region and thighs were performed for nine minutes, beginning approximately 56 and 68 minutes post-injection, respectively. Arterialised blood samples were collected to measure plasma radioactivity and derive the input function for PET data modelling.

### 2.5 PET-image analysis and modelling

Raw [^18^F]FDG-PET/CT images were corrected for attenuation, dead time and decay and reconstructed using the block sequential regularized expectation maximization algorithm (BETA factor 150). Regions of interests (ROIs) for the VL and ASAT were manually delineated in Carimas software, with CT scans serving as anatomical references. Glucose uptake was quantified [33] using the fractional uptake ratio (FUR) method based on arterial input [36].

### 2.6 Biopsy obtainment from skeletal muscle and abdominal subcutaneous adipose tissue

Skeletal muscle (vastus lateralis, VL, n=8 MZ twin pairs) and abdominal subcutaneous adipose tissue (ASAT, n=9 MZ twin pairs) samples were collected in a post-prandial state (1-2 hours) at baseline and post training as previously described[34] at least 48 hours after the last exercise session to ensure no effect of the last exercise session. The biopsies were obtained at the same time from both co-twins, except for one twin pair for whom the biopsies were collected on separate days. VL biopsies were obtained using a suction-modified Bergström technique from a site inferior to trochanter major and anterior to lateral fascia, while ASAT samples were collected from the right side of the lower abdomen via open surgical procedure. All biopsies were immediately flash frozen in liquid nitrogen and stored at –70 °C until homogenization and RNA/DNA co-extraction.

### 2.7 Tissue sample homogenization, RNA and DNA extraction

Frozen VL and ASAT samples were homogenized using a TissueLyser (Qiagen, Hilden, Germany) for 2 × 2 minutes at 25 Hz for VL tissue; and 2 × 1.5 min at 20 Hz for ASAT. RNA and DNA were co-extracted from tissue lysates using AllPrep DNA/RNA/miRNA Universal Kit (Qiagen, Hilden, Germany) following the manufacturer’s protocol. RNA and DNA quality was verified prior to mRNA-sequencing (mRNA-seq) and reduced representation bisulfite sequencing (RRBS) using an Advanced Analytical Fragment Analyzer (Advanced Analytical Technologies, Inc., Ankeny, IA, USA) yielding mean ± SD RQN values of mean 9.11 (SD) 0.25 and mean 7.82 (SD) 0.58 for VL and ASAT RNA, respectively. DNA integrity was inspected visually from gel images and RNA/DNA concentrations were quantified using Qubit®/QuantIT® Fluorometric Quantitation (Life Technologies, Waltham, MA, USA).

### 2.8 RNA Sequencing

Libraries were prepared using the Illumina Stranded mRNA Library Preparation protocol (Illumina, 1000000124518) with the Illumina Stranded mRNA Preparation Ligation Kit (Illumina, Inc, San Diego, California, USA) and 100 ng of total RNA. Library size distribution was assessed with an Advanced Analytical Fragment Analyzer (Advanced Analytical Technologies, Inc., Ankeny, IA, USA) and all the fragments were above 300 bp. RNA concentrations were measured using Qubit®/Quant-IT® Fluorometric Quantitation (Life Technologies, Waltham, MA, USA). Sequencing was performed on an Illumina NovaSeq 6000 S4 v1.5 platform (Illumina, Inc, San Diego, California, USA) with paired-end reads (2 × 150 bp).

#### 2.8.1 RNA-seq data analysis for transcriptome and computational transcription factor (TF) activity

Paired-end reads were aligned to human genome (Homo sapiens GRCh38.109) using Hisat2 (version 2.2.1) and HTSeq (version 0.6.0) in Chipster software. Differential expression analysis was performed using the DESeq2 pipeline (version 1.44.0), with pre-filtering criteria consistent with those applied to methylation data (detailed below) resulting in 17,205 genes for ASAT and 14,355 genes for VL. Quality control included principal component analysis (PCA) on variance-stabilized counts for the 500 most variable genes to identify potential outliers and mitigate skew from extreme count values. No outliers were detected, and sample integrity was verified by low hemoglobin beta (HBB) expression indicating minimal blood contamination and consistent quality across samples. Gene Set Enrichment Analysis (GSEA) was performed using the ReactomePA package (version 1.48.0). For preranked analysis, gsePathway() was applied to all genes ranked by decreasing DESeq2 log2 fold change. For over-representation analysis, enrichPathway() was applied to selected gene subsets, using the tissue-specific background gene list as the universe. For both analyzes, the maximum gene set size was set to 10,000. Transcription Factor (TF) activity was inferred using DecoupleR (version 2.10.0) via the run_ulm function applied to p ≤ 0.05 filtered genes ranked by descending log2 fold change from DESeq2, with a minimum gene set size of five. TF-gene interaction data were retrieved using The Transcription Factor to Gene Target Interaction Database[37] using the get_collectri function from OmnipathR (version 3.12.14). Additionally, TF activity was estimated for Reactome terms by using the above method on Reactome term-specific gene lists and averaged the directional TF activity score, to elucidate TF contributions to enriched pathways. Correlations between Reactome terms and phenotype measures were obtained by performing single-sample gene set enrichment analysis with the GSVA package (version. 2.2.0) using Reactome gene sets, followed by Pearson correlation between the individual Reactome term enrichment scores at baseline or for the change (POST minus PRE) and the phenotype measures. Significant correlations between previously identified Reactome terms and phenotypes were visualized using ComplexHeatmap (version 2.24.0). Individual gene expression change POST – PRE was correlated with log2FC transformed phenotype measure change POST – PRE using Spearman correlation across both twin-pairs. ASAT cell population deconvolution was performed using the CIBERSORT implementation in the IOBR package (version 0.99.0), with the AT22[38] adipose-specific dataset as the signature matrix over 200 permutations. Further deconvolution of macrophage subpopulations was based on the M0, M1 and M2 signatures in the lm22 signature matrix over 200 permutations. The Wilcoxon test was used to determine significantly differing cell fractions. Fiber type proportions were estimated from RNA-seq counts using FibeRtypeR[39]. Although no systematic differences in fiber type composition were observed between groups or time-points, fiber type proportions were included as covariates in differential expression analysis for the VL samples to account for sample-level variation and to minimize potential confounding effects.

### 2.9 Reduced representation bisulfite sequencing (RRBS) for DNA methylome analysis

Libraries were prepared following the Nugen Ovation RRBS Methyl-Seq System protocol (Tecan, M01394) using the Nugen Ovation RRBS Methyl-Seq with TrueMethyl BS (Tecan) and spike-in control of unmethylated lambda DNA (Promega). A starting amount of 100 ng of genomic DNA was used for each library. Library size distribution (fragment sizes were between 229-314 bp) was assessed using the Advanced Analytical Fragment Analyzer (Advanced Analytical Technologies, Inc), and concentrations were determined by fluorometric quantification with Qubit®/Quant-IT® (Life Technologies). Sequencing was performed using NovaSeq 6000 S1 v1.5 with paired-end reads (2 x 100 bp) across two lanes.

#### 2.9.1 RRBS data analysis

Reduced representation bisulfite sequencing (RRBS) data were analyzed using a modified MethylKit pipeline (R package, version 1.30.0). Coverage normalization was performed with the normalizeCoverage function. CpG sites were retained if present in at least 6 samples per time × condition within ASAT or VL samples, and had a minimum coverage of 10 reads resulting in total of 1,600, 783 sites analyzed for ASAT and 1,843,031 for the VL. Initial mixed-effects modeling of methylation levels using the formula: methylation_levels ∼ time_point * twin_status + (1 | FP) + (1 | twin_id) with lmer function (lme4, version 1.1-35.5) revealed substantial multi co-linearity, preventing model convergence. Differentially methylated regions (DMRs) were detected by aggregating methylated and un-methylated counts into 1,000 bp tiles using tileMethCounts, followed by differential methylation analysis using the same approach as used for DMPs. Annotation of DMPs and DMRs to the GRCh38/hg38 genome assembly was performed with the annotatr package (version 1.30.0), generating gene and CpG annotation data frames via build_annotations and genomic coordinates with methylation sites. GSEA for DMRs was performed using the ReactomePA package (version 1.48.0), with the gsePathway function applied to DMRs ranked by descending diff.meth change from calculateDiffMeth function in MethylKit described above. As suggested above, using FibeRtypeR, fiber proportions were included as covariates in DMP and DMR analysis for the VL samples to account for sample level variability and reduce potential confounding effects on differential methylation analysis. Due to the modest magnitudes of gene expression and DNA methylation changes typically observed in resting human skeletal muscle and adipose tissue post training, combined with substantial inter individual variability and the restricted sample size inherent to MZ twin study designs, FDR correction proved overly conservative and eliminated many biologically relevant signals. Therefore, consistent with a prior multi-omics analyzes in BMI discordant MZ twin cohorts[6], we applied nominal p ≤ 0.05 together with biologically justified effect size thresholds (≥ ±20 percent change for gene expression and ≥ ±5 percent methylation difference) to retain physiologically meaningful signals. The effect size thresholds used here were chosen to reflect the known dynamic ranges of each omics layer in human exercise studies, where DNA methylation changes of approximately 5-20 percent are typical and biologically relevant in skeletal muscle after training[40] or inactivity[41], whereas the majority of resting, post training gene expression changes lie within approximately 1.2-1.5 fold (20–50%)[24,42]. Applying these biologically grounded thresholds alongside nominal p value testing helps avoid underestimation of integrative methylation-expression overlap. Furthermore, applying the same significance criteria to both skeletal muscle and ASAT enabled direct cross-tissue comparison and ensured that candidate features were prioritized based on both statistical support and effect magnitude.

Differentially methylated and differentially expressed genes (DmDEGs) were manually overlapped by mapping DMRs to DEGs by their ENSEMBL ids. For genes with more than one significant DMR in a given contrast, we assigned each DMR a weight proportional to the absolute methylation difference, the negative base-10 logarithm of the nominal p-value, and an inverse distance-to-transcription-start-site term, defined as 1/(1 + Δ*i*/1000), where Δ*i* is the distance in base pairs from the middle of the DMR to the transcriptional start site. The gene-level weighted mean methylation difference was then calculated as the weighted average of DMR methylation differences across all significant DMRs assigned to that gene. In parallel, a weighted gene score was computed as the sum of signed DMR weights to prioritize genes with stronger aggregate methylation evidence.

### 2.10 Statistical analyzes

Physiological data were analyzed using SAS software (version 9.4; SAS Institute, Cary, NC, USA). Two-sided tests were applied, and p-values ≤ 0.05 were considered statistically significant. Normality was assessed visually using Q-Q plots, histograms and studentized residuals. Analyzes employed a linear mixed-effects model for repeated measures with a compound symmetry covariance structure. The model included twin as the statistical unit, time (PRE and POST intervention), twin group (leaner vs. heavier co-twin), and their interaction (time×group). Restricted maximum likelihood estimation was used allowing inclusion of participants with missing data. When a significant time × group interaction was detected, the same model was applied to assess within-group effects over time. Baseline differences between co-twins were estimated using pre-intervention data within the same model. RNA-seq and RRBS data were analyzed using R (version 4.5.0) using the packages and functions specified above, and packages used ubiquitously across both omics analysis include tidyverse (version 2.0.0), ggplot2 (version 3.5.1), ggrepel (version 0.9.6), patchwork (version 1.3.0), stringer (version 1.5.1), parallel (version 4.4.0), annotatr (version 1.30.0), annotationDBI (version 1.66.0) and org.Hs.eg.db (version 3.19.1).

## 3 RESULTS

### 3.1 BMI-Dependent Baseline Differences Between Heavier and Leaner co-twins with Similar Training Responses in Cardiorespiratory Fitness without Altering Whole-Body Fat Percentage

At baseline, heavier co-twins exhibited greater whole-body fat percentage by 33% (ES: 10.15%, 95% CI: [5.57;14.74], *p* < 0.001) as well as lower cardiorespiratory fitness (VO_2peak_) by –27% (ES: –6.8 ml/kg/min, 95% CI: [-10.66; –2.93], *p* = 0.003) and whole-body insulin sensitivity (M-value) by – 63% (ES: –14.48 µmol/kg/min, 95% CI: [-23.7;-5.3], *p* = 0.007) compared with leaner co-twins (Table 1). Following the training intervention, VO_2peak_ and M-value improved by 9.4% (ES: 2.7 ml/kg/min, 95 % CI [1.3, 4.2], *p* = 0.001) and 29.4% (ES: 8.9 µmol/kg/min, 95 % CI [1.5; 16.3], *p* = 0.022), respectively, with similar training effects in both twin groups. Whole-body fat percentage remained unchanged with training (*p* = 0.370) in both groups. Visceral fat mass decreased only in the heavier co-twins by 0.37 kg, (95 % CI: [0.045; 0.70], *p* = 0.029).

**Table 1.** Baseline and Post-Exercise Training Participant Characteristics. Values are model-based means with 95% confidence intervals.

|  | Leaner co-twins |  | Heavier co-twins |  | p-value |  |  |
| --- | --- | --- | --- | --- | --- | --- | --- |
|  | Pre (n=12) | Post (n=10) | Pre (n=12) | Post (n=11) | Baseline | Time | Time*group |
| Sex | 8 female / 4 male pairs |  |  |  |  |  |  |
| Age | 40.4 [37.5;43.4] |  | 40.4 [37.5;43.4] |  |  |  |  |
| Anthropometrics |  |  |  |  |  |  |  |
| Weight (kg) | 86.4 [72.4;100.4] | 86.9 [72.6;101.2] | 108.7 [94.2;123.3] | 108.0 [93.1;122.9] | 0.001 | 0.948 | 0.374 |
| BMI (kg/m²) | 29.1 [25.2;33.0] | 29.3 [25.3;33.2] | 36.7 [32.7;40.7] | 36.4 [32.4;40.4] | 0.0006 | 0.921 | 0.407 |
| Whole-body fat (%) | 30.4 [21.3;39.6] | 29.5 [20.3;38.7] | 40.6 [36.5;44.7] | 40.0 [35.9;44.1] | 0.0005 | 0.370 | 0.718 |
| Lean mass (kg) | 33.1 [30.0;36.3] | 33.9 [30.6;37.2] | 35.9 [31.9;39.8] | 36.2 [32.1;40.3] | 0.003 | 0.140 | 0.102 |
| Lean mass (kg) | 33.1 [30.0;36.3] | 33.9 [30.6;37.2] | 35.9 [31.9;39.8] | 36.2 [32.1;40.3] | 0.003 | 0.140 | 0.102 |
| Visceral fat mass (kg) | 3.38 [2.13;4.64]† | 3.22 [2.03;4.42] | 5.83 [4.74;6.93]§ | 5.46 [4.42;6.50]‡ | 0.002 | 0.067 | 0.280 |
| Glucose profile |  |  |  |  |  |  |  |
| Fasting glucose (mmol/l) | 5.5 [5.2;5.7] | 5.5 [5.2;5.7] | 5.7 [5.4;5.9] | 5.8 [5.6;6.1] | 0.388 | 0.367 | 0.418 |
| Fasting insulin (mU/l) | 6.6 [5.1;8.7] | 6.3 [4.3;9.3] | 11.1 [8.7;14.2] | 9.9 [6.9;14.1] | 0.006 | 0.502 | 0.711 |
| HbA1c (mmol/mol) | 34.9 [32.9;36.9] | 34.7 [32.2;37.1] | 36.5 [35.0;38.0] | 36.0 [34.2;37.7] | 0.047 | 0.581 | 0.675 |
| M-value (μmol/kg/min) | 37.5 [28.0;47.0]† | 46.9 [31.7;62.1]‡ | 23.0 [16.1;29.9]† | 31.4 [20.4;42.3]§ | 0.007 | 0.022 | 0.82 |
| Others |  |  |  |  |  |  |  |
| VO<sub>2peak</sub> (ml/kg/min) | 32.4 [26.9;37.8] | 35.1 [29.9;40.2] | 25.6 [23.2;28.0] | 28.3 [26.1;30.6] | 0.003 | 0.001 | 0.935 |
| hs-CRP (mg/l) | 0.81 [0.41;1.61]‡ | 0.56 [0.21;1.47]§ | 1.42<br>[0.75;2.70]‡ | 1.14<br>[0.46;2.85]¶ | 0.005 | 0.296 | 0.450 |
| **Average energy intake (kCal)** | 1950 [1657;2242] | 2145 [1861;2429] | 2085 [1808;2362] | 2198 [1921;2475] | 0.306 | 0.076 | 0.574 |

*P*-value for **baseline** indicates differences between heavier and leaner co-twins before the exercise training intervention. *P*-value for **time** reflects changes from baseline (PRE) to after (POST) training across all participants. *P*-value for **time × group interaction** represents differences in training-induced changes between heavier and leaner co-twins. † *n* = 11; ‡ *n* = 10; § *n* = 9; ¶ *n* = 8.
**Abbreviations:** VO<sub>2peak</sub> = cardiorespiratory fitness; hs-CRP = high-sensitivity C-reactive protein; M-value = whole-body insulin sensitivity; HbA1c = glycated hemoglobin; GU = Glucose uptake For time and time × group analyzes, hs-CRP and fasting insulin were log-transformed, for baseline comparisons, visceral fat mass, hs-CRP and HbA1c, were log-transformed; lean mass was square-root transformed. hs-CRP was measured in a post-prandial state prior to the [<sup>11</sup>C]PK11195 scan[34]

### 3.2 Exercise Training Has No Effect on Insulin-Stimulated GU But Reduces Transcriptomic Divergence in ASAT Between the Heavier and Leaner Monozygotic Co-Twins

Given the higher insulin-stimulated ASAT GU in leaner co-twins compared with their heavier co-twins (*p* < 0.01; Figure 2A & B) and the absence of a training effect on ASAT glucose uptake (*p* = 0.440; Figure 2B), we next examined whether ASAT displayed an obesity-associated molecular profile and whether this was modified by exercise training. At baseline, 1625 genes were differentially expressed in heavier compared with leaner co-twins in ASAT (870 upregulated, 755 downregulated; Figure 2C). Exercise training elicited a substantially larger transcriptional response in heavier co-twins (840 DEGs: 458 upregulated, 382 downregulated; Figure 2D) than in leaner co-twins (252 DEGs: 111 upregulated, 141 downregulated; Figure 2E). After 6 months of training, transcriptomic profiles converged, with the number of DEGs between co-twins decreasing by ∼36% (post-intervention: 1035 DEGs, 634 upregulated, 401 downregulated; Figure 2F), although 381 DEGs remained significantly different. Only 24 training-induced genes were regulated in the same direction in both groups: 7 downregulated (Figure 2G) and 17 upregulated (Figure 2H), indicating that exercise induced largely distinct transcriptional reprogramming depending on baseline adiposity. The full DEG-lists are provided in Supplementary File 1. To quantify convergence, Euclidean distance between co-twins PCA space was assessed for all baseline DEGs between time points. Training reduced this distance by –5.2 units (95% CI: –11.1 to 0.6, p = 0.053; Figure 2I), suggesting that exercise training shifted the transcriptome of the heavier co-twins towards that of their leaner counterparts.

**Figure 2.**
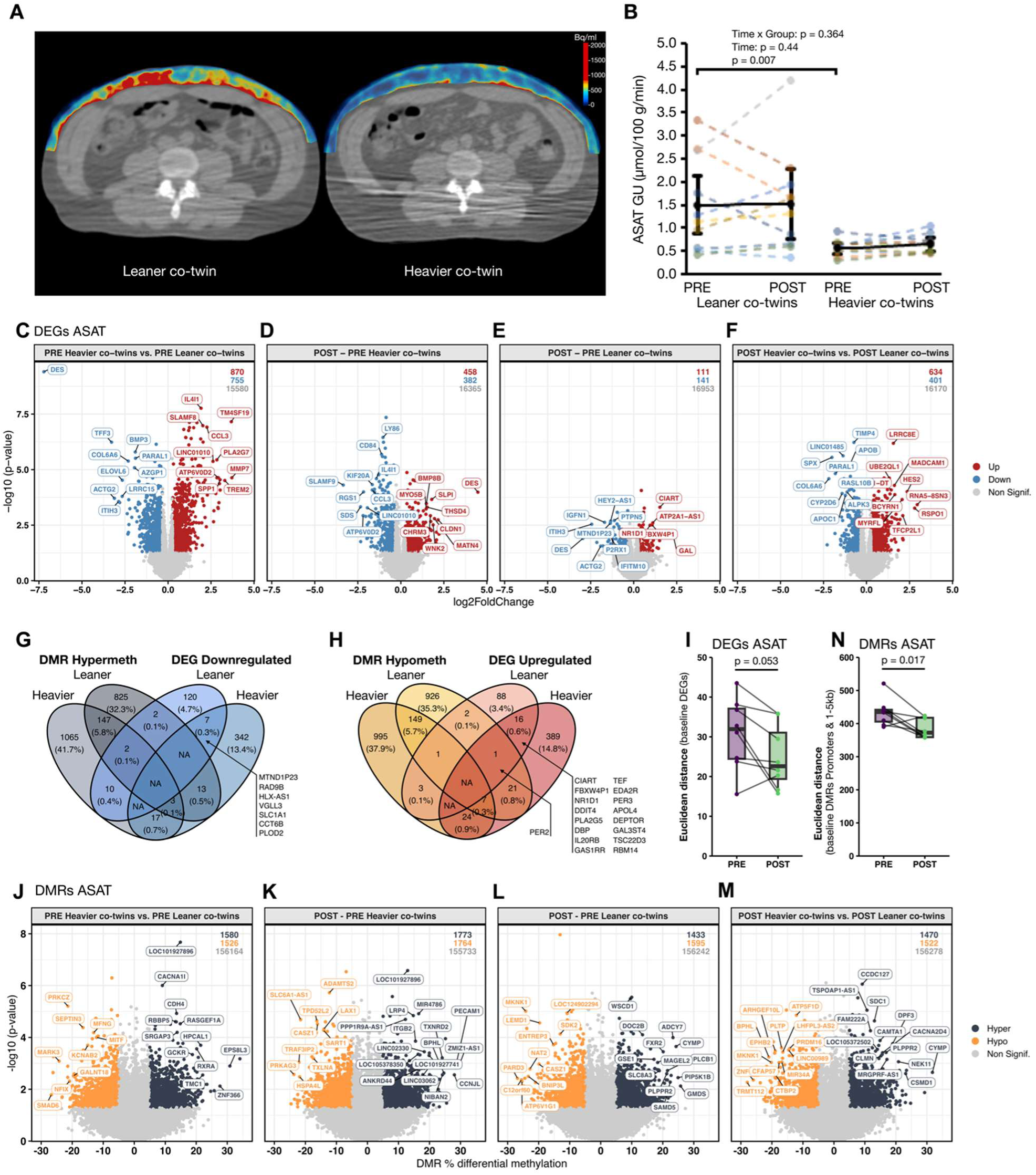
Insulin-stimulated glucose uptake (GU), gene expression and DNA methylation in abdominal subcutaneous adipose tissue (ASAT) A) Representative [^18^F]FDG-PET/CT-images of a leaner and heavier co-twin at baseline in ASAT. B) Insulin-stimulated glucose uptake in ASAT in leaner and heavier co-twins at PRE and POST intervention. Bolded line depicts model based mean with 95% confidence intervals. Co-twins share the same colour in the figure. C) Volcano plot of differentially expressed genes (DEGs) between heavier and leaner co-twins before exercise training intervention (PRE). D) Volcano plot of DEGs in response to exercise training intervention in heavier and E) leaner co-twins. F) Volcano plot of DEGs between heavier and leaner co-twins after exercise training intervention (POST). In C-F, the cut-off for statistical significance was set at nominal p ≤ 0.05 with ≥20% difference between co-twins (C&F) or ≥20% change following exercise training (D&E). Top DEGs ranked by XIAO score are annotated in the volcano plots. Displayed values indicate the number of upregulated, downregulated, and non-significant genes per comparison. G) Venn diagram of overlap between downregulated DEGs and all differentially hypermethylated regions (DMRs). H) Venn diagram of overlap between upregulated DEGs and all hypomethylated DMRs. I) Euclidean distance in principal component space based on baseline DEGs depicted as boxplots (median, IQR) showing within twin pair divergence at baseline (PRE) and after exercise training (POST); statistical significance was tested by paired t-test. J) Volcano plot of all DMRs between heavier and leaner co-twins before exercise training intervention (PRE). K) Volcano plot of all DMRs in response to exercise training intervention in heavier and L) leaner co-twins. M) Volcano plot of DMRs between heavier and leaner co-twins after exercise training intervention (POST). In I-L, the cut-off for the statistical significance was set at nominal p ≤ 0.05 with either ≥5% difference between heavier and leaner co-twins (I&L) or ≥5% change following exercise training (K&L). Top DMRs ranked by XIAO score are annotated in the volcano plots. Displayed values indicate the number of hypermethylated, hypomethylated, and non-significant regions in each comparison. N) Euclidean distance in principal component space based on baseline DMRs in promoters and 1-5kb from transcriptional start site, depicted as boxplots (median, IQR) showing within twin pair divergence at baseline (PRE) and after exercise training (POST); statistical significance was tested by paired t-test.

### 3.3 Differentially Methylated Regions Reveal Pronounced Training Effects in ASAT of Heavier Co-Twins

To determine whether the transcriptional remodeling in ASAT was accompanied by epigenetic reprogramming, we next analyzed differential DNA methylation between co-twins at baseline and in response to training. At baseline, the heavier co-twins exhibited 3,106 DMRs compared with the leaner co-twins, including 1,580 hypermethylated and 1,526 hypomethylated regions across all annotated regions (Figure 2J). Heavier co-twins also demonstrated a greater training-induced epigenetic reprogramming (+17%) with 3,537 DMRs versus 3,028 in leaner co-twins (Figures 2K & L). Only 451 training-evoked DMRs overlapped between groups, of which 157 DMRs were hypomethylated (Figure 2G) and 152 DMRs hypermethylated (Figure 2H) in both groups, indicating that the majority of exercise responsive DMRs are either uniquely regulated or discordantly methylated between heavier and leaner co-twins. After training, 2,992 DMRs were detected between heavier and leaner co-twins (Figure 2M), however, only 115 were retained from baseline (Supplemental Figure 2A), with 23 remaining hypomethylated and 25 hypermethylated (Supplemental Figure 2B). These findings indicated a more pronounced effect on DNA methylation in heavier co-twins and distinct DNA methylation signatures in ASAT between groups. Full DMR-lists are provided in Supplemental File 2. Focusing on DMRs in regulatory regions (promoters and 1-5 kB upstream of the transcriptional starting site (TSS)) with a high potential to regulate gene expression, 947 baseline regulatory DMRs were identified (473 hypermethylated, 474 hypomethylated in heavier co-twins; Supplemental Figure 2C). Training induced hypermethylation of 616 and hypomethylation of 504 regulatory DMRs in heavier co-twins (Supplemental Figure 2D), and 470 hypermethylated and 506 hypomethylated regulatory DMRs in leaner co-twins (Supplemental Figure 2E), with 420 hypermethylated and 508 hypomethylated regulatory DMRs remaining after training (Supplemental Figure 2F). Of these, 57 training-induced regulatory DMRs overlapped between groups (26 hypermethylated, 31 hypomethylated; Supplemental Figure 2G & H). Euclidean distance between co-twins in the PCA space, based on baseline regulatory DMRs, showed a significant convergence (Figure 2N; p = 0.017) following the training intervention. Together, these findings show that exercise induces extensive epigenetic remodeling in ASAT, with a strong convergence occurring at promoter and 1-5kb from TSS regulatory regions.

### 3.4 Integration of DNA Methylation and Gene Expression Reveals Distinct Exercise Responses in ASAT of heavier co-twins

Next, we overlapped the Differentially Methylated regions with the Differentially Expressed Genes, termed DmDEGs, to identify DEGs potentially regulated by DNA methylation. To avoid missing biologically relevant DmDEGs, DEGs were overlapped with DMRs associated with exons, introns and 5’UTRs, representing an inclusive gene-associated annotation that encompasses promoter and 1-5 kb regions. Promoter and 1-5 kb from TSS associated DMRs were then additionally analyzed as a distinct regulatory subset. At baseline, 144 DmDEGs were identified, regulated by 175 DMRs with 23 DEGs associated with multiple DMRs (Figure 3A, full annotation provided in Supplementary File 3.1). Of these DmDEGs, 43 were hypomethylated-upregulated and 35 were hypermethylated-downregulated in heavier versus leaner co-twins (Figure 3A). Across all genomic regions, intronic DMRs contributed the greatest overlap (21 hypomethylated-upregulated; 19 hypermethylated-downregulated; Supplementary File 3.2, likely reflecting their higher absolute abundance. The hypomethylated-upregulated DmDEGs grouped predominantly to immune system and cell cycle pathways. Key immune related examples included *ITGB5*, *PDCD1*, *ITGB2*, *KCNAB2*, *BATF*, *PLAUR* and *ARPC1B*. Cell cycle related examples included *NINL*, *RMI2* and *TPX2*. Similarly, the hypermethylated-downregulated DmDEGs were enriched for lipid metabolism and small molecule transport. Representative genes included *FASN*, *PPARA*, *ACACA*, *LPIN1* and *NPAS2*, alongside transporters such as *SLC38A3*, *SLC2A6*, and *SLC4A4*. *PRDM16* (adipocyte browning) was hypermethylated-downregulated. Given the extensive gene list, full mapping and genomic region annotation are provided in Supplementary File 3.1, and weighted mean methylation difference per gene in Supplementary File 3.2.

**Figure 3.**
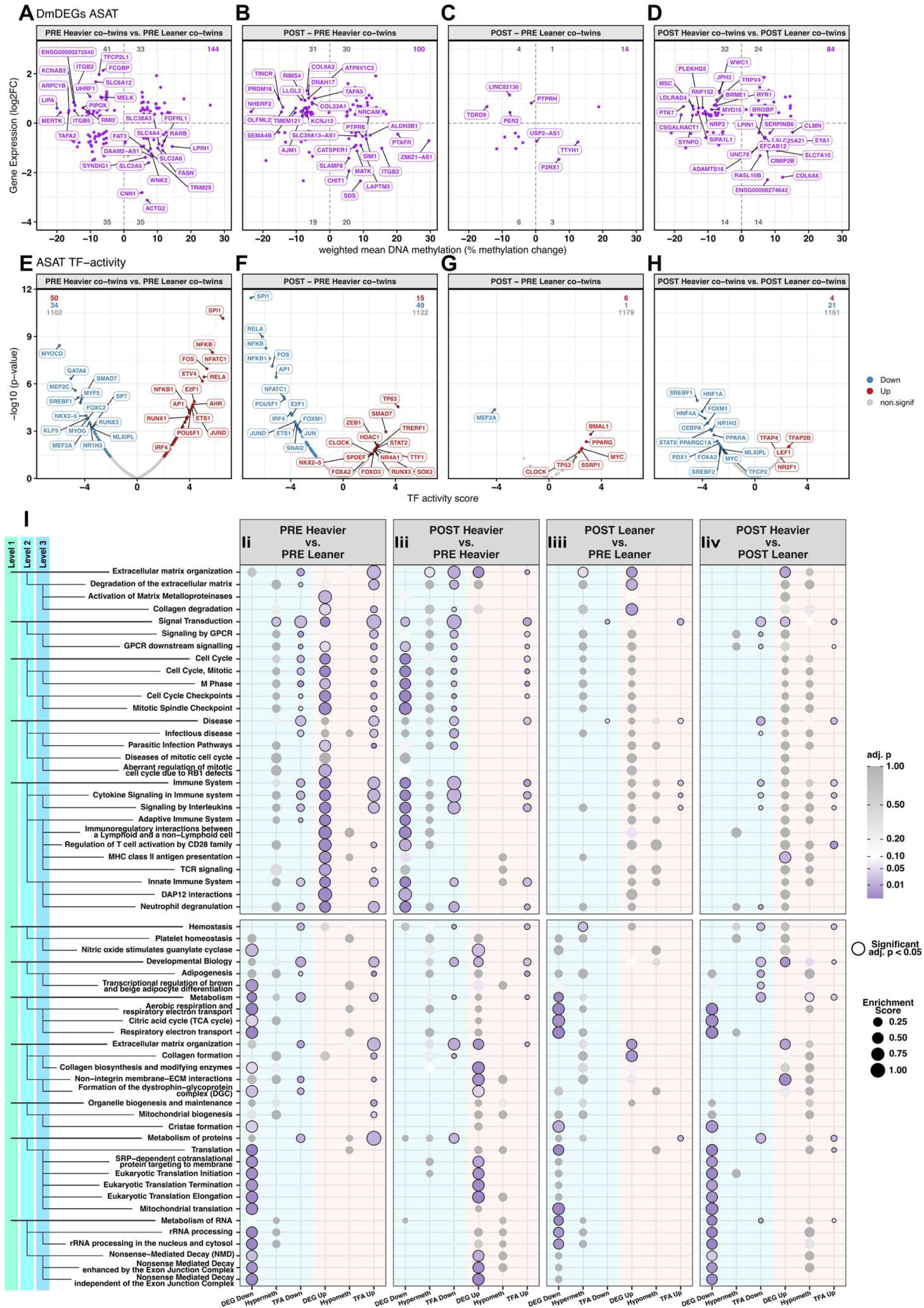
Identification of differentially methylated and expressed genes (DmDEGs), transcription factor activity (TFA), and Reactome pathway enrichment analysis in abdominal subcutaneous adipose tissue (ASAT) A) Volcano plot of differentially methylated and expressed genes (DmDEGs) between heavier and leaner co-twins before exercise training intervention (PRE). B) Volcano plot of DmDEGs in response to exercise training intervention in heavier and C) leaner co-twins. D) Volcano plot of DmDEGs between heavier and leaner co-twins after exercise training intervention (POST). In A-D, for genes with more than one significant DMR in a given contrast, we assigned each DMR a weight proportional to the absolute methylation difference, the negative base-10 logarithm of the nominal p-value, and an inverse distance-to-transcription-start-site term, and calculated the weighted mean methylation difference. E) U-plot depicting the difference in transcription factor activity ((TFA) estimated from gene expression data using decouplR)) between heavier and leaner co-twins before intervention (PRE). F) U-plot depicting the difference in TFA in response to exercise training in heavier and G) leaner co-twins. H) U-plot depicting the difference in TFA between heavier and leaner co-twins after the exercise intervention (POST). Ii) A bubble plot showing the top up– and downregulated Reactome pathways identified by differential gene expression between heavier and leaner co-twins before exercise training intervention (PRE). For the top enriched terms identified by gene expression, the role of DNA methylation and TFA were also assessed. Iii) A bubble plot showing the effect of exercise training on the gene expression, DNA methylation and TFA for the top enriched terms identified in (Ii) in heavier and Iiii) leaner co-twins. Iiv) A bubble plot showing the difference in gene expression, DNA methylation and TFA between heavier and leaner co-twins identified at (Ii) after the exercise training intervention (POST). The colour and the size of the each bubble depicts the degree of the statistical significance and enrichment score, respectively. Bubbles circled with black solid line depict statistically significant (adj. p ≤ 0.05) terms in each comparison.

Training-induced changes (POST vs. PRE) in heavier co-twins yielded 100 DmDEGs (Figure 3B). Of these genes, 31 were hypomethylated-upregulated genes representing ECM remodeling (e.g., *FBLN1*, *COL23A1*, *COL9A3*), signal transduction (e.g., *GNAO1*, *ITPR3*, *RAMP3* and *PAK4*), and adipocyte browning (*PRDM16*). Additionally, 20 DmDEGs were hypermethylated-downregulated, enriched for inflammatory and lysosomal pathways (e.g., *CCL2*, *MATK*, *SLAMF8*, *CHIT1, PTPRE*, *PLA2G15*, *LAPTM5*, *RAB7B*). In response to training, 14 DmDEGs were identified, in leaner co-twins (4 hypomethylated-upregulated and 3 hypermethylated-downregulated; Figure 3C), indicating a more modest response relative to heavier co-twins. Thus, exercise training reduced the number of DmDEGs between co-twins, with 84 identified post-training (32 hypomethylated-upregulated; 14 hypermethylated-downregulated; Figure 3D). Intronic methylation again exhibited the highest degree of overlap with gene expression (Supplementary file 3.1). Importantly, no DmDEGs overlapped between baseline and post-training, indicating distinct regulatory responses. A full list of DmDEGs is provided in Supplementary Files 3.1 & 3.2.

### 3.5 Exercise Training Normalizes Transcription Factor Activity in ASAT, With Greater Regulatory Shifts in Heavier Co-Twins

We observed a large number of differentially expressed genes (DEGs) and differentially methylated regions (DMRs) in ASAT; however, only approximately 2-5 % of genes showed both differential methylation and differential expression, suggesting a notable role of a second regulatory mechanism in addition to DNA methylation. To explore this discrepancy, transcription factor (TF) activity was inferred from RNA-seq data to assess regulatory influences on DEGs. At baseline, 50 and 34 TFs were more and less active, respectively, in heavier compared to leaner co-twins (Figure 3E). Training effects were more pronounced in the heavier co-twins, with 15 TFs increasing and 49 TFs decreasing activity (Figure 3F), compared with leaner co-twins where only six TFs (*BMAL1*, *CLOCK*, *MYC*, *PPARG*, *SSRP1* and *TP53*) increased and one (*MEF2A*) decreased activity (Figure 3G). Importantly, exercise training normalized the activity of many TFs that were elevated at baseline in heavier co-twins, normalizing activity toward leaner co-twins (Figure 3E & H). *LEF1*, a mediator of Wnt signaling implicated in adipose tissue remodeling, was the only TF that was more active in heavier co-twins at baseline and that remained more active after training. In contrast, *SREBF1*, *SREBF2*, *MLXIPL*, *HNF4A*, *NR1H3*, *NR2F2* & *NR4A1* were less active in heavier co-twins at baseline, and remained less active after 6 months of training. Several of these TFs are key regulators of lipid and glucose metabolism, including de novo lipogenesis, sterol sensing, and nutrient-responsive transcription, indicating persistent suppression of a metabolic regulatory network in the heavier co-twins despite exercise training. The entire list of differentially active TFs is provided in Supplementary File 4.

### 3.6 Pathway-Level Reversal of Obesity-Associated Transcriptomic Signatures in ASAT Following Exercise Training

To elucidate the biological relevance of DEGs associated with obesity and modulated by exercise training, we performed Reactome pathway enrichment analysis. At baseline, heavier co-twins exhibited upregulation of pathways related to the immune system, mitosis, cell cycle and collagen degradation by matrix metalloproteinases, indicating a transcriptional signature related to increased adipose tissue inflammation, cellular proliferation and extracellular matrix breakdown (Figure 3Ii). These transcriptional differences may be partially explained by significantly elevated macrophage content in ASAT biopsies (*p* = 0.026) from heavier co-twins (Supplementary Figure 6). Notably, transcriptional upregulation of these pathways was not accompanied with differential methylation at pathway level. Instead, transcription factor (TF) activity appeared to drive these changes (Figure 3Ii).

Conversely, downregulated pathways in heavier co-twins were related to aerobic metabolism, including electron transport chain and citric acid cycle, suggesting impaired aerobic metabolism and mitochondrial function (Figure 3Ii). In addition, pathways governing transcriptional regulation of brown and beige adipocyte differentiation, collagen biosynthesis, protein translation, rRNA processing, and nonsense-mediated decay were downregulated in heavier co-twins compared with their leaner co-twins (Figure 3Ii). These transcriptional reductions were also not explained by hypermethylation at pathway level, although several were associated with reduced TF activity (Figure 3Ii).

Next, we investigated whether exercise training modified the ASAT Reactome pathways that differed at baseline between heavier and leaner co-twins. Exercise training attenuated the obesity-associated transcriptional activation of inflammatory and proliferative pathways in heavier co-twins resulting in non-significant differences in these pathways after the intervention (Figure 3Iii & 3Iiv), in line with the observed transcriptional and epigenetic convergence (Figure 2G & H). The training-induced downregulation of these pathways was accompanied by a statistically non-significant increase in DNA hypermethylation at pathway level and a statistically significant reduction in TF activity (Figure 3Iii), indicating that transcriptional regulatory mechanisms may contribute more substantially than DNA methylation to the suppression of inflammatory and proliferative gene expression in heavier co-twins. Furthermore, exercise training restored the obesity-associated downregulation of pathways related to collagen biosynthesis, protein translation and nonsense-mediated decay pathways in heavier co-twins, and these changes were not accompanied by changes in DNA methylation or TF activity at the pathway level (Figure 3Iii). Interestingly, exercise training upregulated genes involved in extracellular matrix (ECM) organization in both co-twin groups, and this effect was not associated with increased TF activity or DNA hypomethylation (Figure 3Iii & 3Iiii). The top 10 up– and downregulated Reactome pathways induced by exercise training independent of baseline differences between co-twin groups, for both DEGs and DMRs in ASAT of heavier and leaner co-twins are shown in Supplementary Figures 4 and 5, respectively.

### 3.7 Associating Adipose Tissue Transcriptomic Pathways to Insulin Sensitivity and Inflammation

We next examined whether the top Reactome pathways enriched for differentially expressed genes (DEGs) between the co-twins at baseline were directly associated with key metabolic, systemic inflammation and physiological measures. To achieve this, we calculated the Reactome term enrichment for each co-twin and both time-points individually and correlated the per sample enrichment score with the whole-body insulin sensitivity (M-value), insulin-stimulated glucose uptake in ASAT, systemic inflammation (hs-CRP), visceral fat mass and BMI. Due to the small sample size, the co-twin groups were pooled to avoid false positive and negative correlations. At baseline, pathways related to the immune system, cell division and extracellular matrix organization correlated negatively with adipose tissue glucose uptake and whole-body insulin sensitivity (M-value; Figure 4A), whereas pathways associated with transcriptional regulation of brown and beige adipocyte differentiation, mitochondrial biogenesis, RNA metabolism and aerobic respiration showed positive correlations with these measures (Figure 4C). Furthermore, immune function and extracellular matrix pathways showed positive associations with BMI and visceral fat mass (Figure 4A), whereas aerobic respiration, transcriptional regulation of brown and beige adipocyte differentiation and RNA metabolism showed negative associations with these measures (Figure 4C). In addition, several immune system pathways correlated positively with hs-CRP (Figure 4A), while transcriptional regulation of brown and beige adipocyte differentiation, mitochondrial biogenesis and metabolism of RNA correlated negatively with hs-CRP (Figure 4C). In contrast, training-induced changes in pathways related to immune function, cell division, aerobic respiration and citric acid cycle did not display systematic correlations with changes in adipose tissue glucose uptake, whole-body insulin sensitivity, or with visceral fat mass when both co-twin groups were analyzed together (Figure 4B & D). Interestingly, the change in the expression of genes involved in translation and RNA processing correlated positively with the change in ASAT glucose uptake. (Figure 4B).

**Figure 4.**
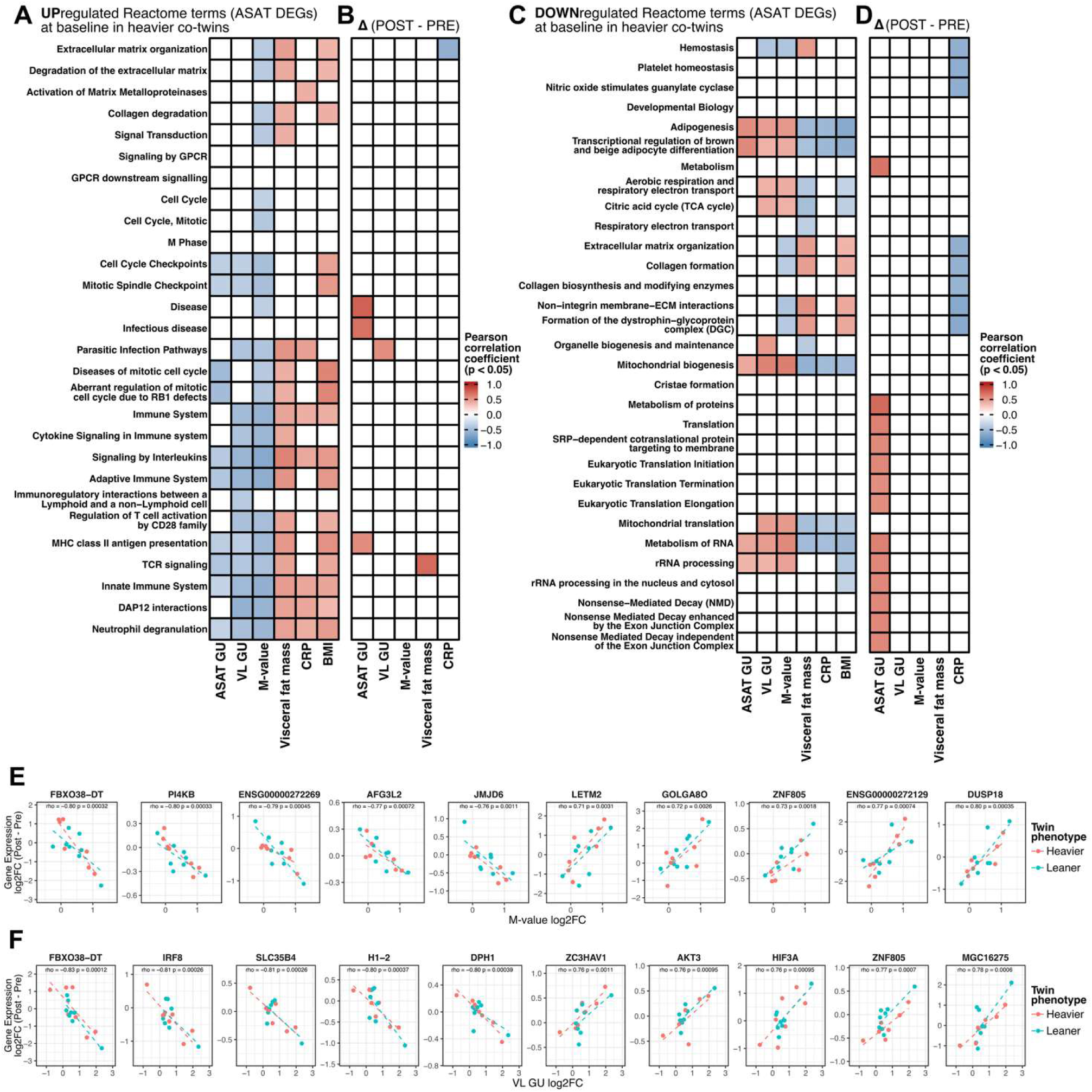
Correlations between gene expression in ASAT and key phenotype measures. A) Correlation heatmap showing Pearson correlations between Reactome terms (upregulated at gene expression level in heavier compared with leaner co-twins before intervention) and key phenotype outcome measures (insulin-stimulated glucose uptake (GU) in abdominal subcutaneous adipose tissue (ASAT) and vastus lateralis (VL), whole-body insulin sensitivity (M-value), visceral fat mass, BMI and high-sensitive C-reactive protein (CRP)). B) Correlation heatmap showing Pearson correlations between the exercise training intervention induced change (“Δ post – pre”) in reactome term enrichment at gene expression level and the change (Δ post – pre) in key phenotype outcome measures. C) Correlation heatmap showing Pearson correlations between reactome terms (downregulated at gene expression level in heavier compared with leaner co-twins before intervention) and key phenotype outcome measures. D) Correlation heatmap showing Pearson correlations between the exercise training intervention induced change (“Δ post – pre”) in Reactome term enrichment at gene expression level and the change (Δ post – pre) in key phenotype outcome measures. Scatterplots depicting the top 5 positively and top 5 negatively correlated (Spearman) individual genes with E) M-value and F) VL GU. In A-D) Only significant correlations (nominal p ≤ 0.05) are colored in the heat map. The colour intensity depicts the correlation coefficient value. Co-twin groups were pooled for the Δ post – pre correlation analyzes (A-E).

### 3.8 Linking Adipose Tissue Transcriptome to Skeletal Muscle and Whole-Body Insulin Sensitivity

Interestingly, baseline correlations between ASAT Reactome pathways and systemic metabolic measures revealed that immune system and cell division pathways were negatively associated with skeletal muscle glucose uptake and whole-body insulin sensitivity (M-value), whereas transcriptional regulation of brown and beige adipocyte differentiation, aerobic respiration and RNA metabolism were positively associated with these measures (Figure 4A & C). These findings prompted us to investigate whether exercise-induced changes in ASAT gene expression were linked to improvements in skeletal muscle and whole-body insulin sensitivity, suggestive of cross-tissue interactions. The ASAT genes most strongly negatively associated with changes in whole body insulin sensitivity (M-value) included *FBXO38-DT*, *PI4KB*, *AFG3L2, ENSG00000272269* and *JMJD6*, while *LETM2*, *GOLGA8O*, *ZNF805, ENSG00000272129* and *DUSP18* showed the strongest positive associations (Figure 4E). The five ASAT genes demonstrating the strongest negative correlations with skeletal muscle glucose uptake were *FBXO38-DT*, *IRF8*, *SLC35B4*, *H1-2* and *DPH1*, whereas the strongest positive correlations were observed for *ZC3HAV1*, *AKT3*, *HIF3A*, *ZNF805* and *MGC16275* (Figure 4F).

### 3.9 Exercise Training Improves Insulin-Stimulated Glucose Uptake in Skeletal Muscle – Modest Transcriptomic Adaptations in Skeletal Muscle Compared to ASAT

Leaner co-twins had higher insulin-stimulated GU in VL compared with their heavier co-twins (p<0.01, Figure 5A & B) while exercise training improved VL GU similarly in both twin groups (Time: p<0.05, Time x group, p=0.672; Figure 5B). However, transcription profiles in VL were more similar between co-twin groups than in ASAT, with 315 DEGs: 213 upregulated and 102 downregulated in heavier compared with their leaner co-twins at baseline (Figure 5C). Exercise training, with the biopsies obtained at rest, yielded 362 DEGs in heavier co-twins and 480 in leaner co-twins (Figure 5D & E). In heavier co-twins, 154 genes were downregulated and 208 were upregulated post-training (Figure 5D, whereas leaner co-twins showed 153 downregulated and 327 upregulated DEGs (Figure 5E). Post-training heavier co-twins exhibited 300 upregulated and 284 downregulated genes compared with leaner co-twins, only 44 DEGs overlapped with baseline DEGs (Figures 5F). Overlap between training-evoked transcriptional responses was minimal: only 10 DEGs were downregulated in both groups (Figure 5G), and 23 DEGs were upregulated in both (Figure 5H). Over-representation analysis indicated that the gene sets unique to each group mapped to mostly distinct Reactome pathways (Supplemental File 5) suggesting group-specific transcriptional adaptations in mainly different pathways. Euclidean distance between co-twins, calculated in the principal component space using baseline DEGs, demonstrated that transcriptomic divergence between co-twins was less affected by training in skeletal muscle than in ASAT, with ASAT differences being approximately three-fold greater at baseline (Figures 2I, 5I). Complete DEG-lists are provided in Supplementary File 1.

**Figure 5.**
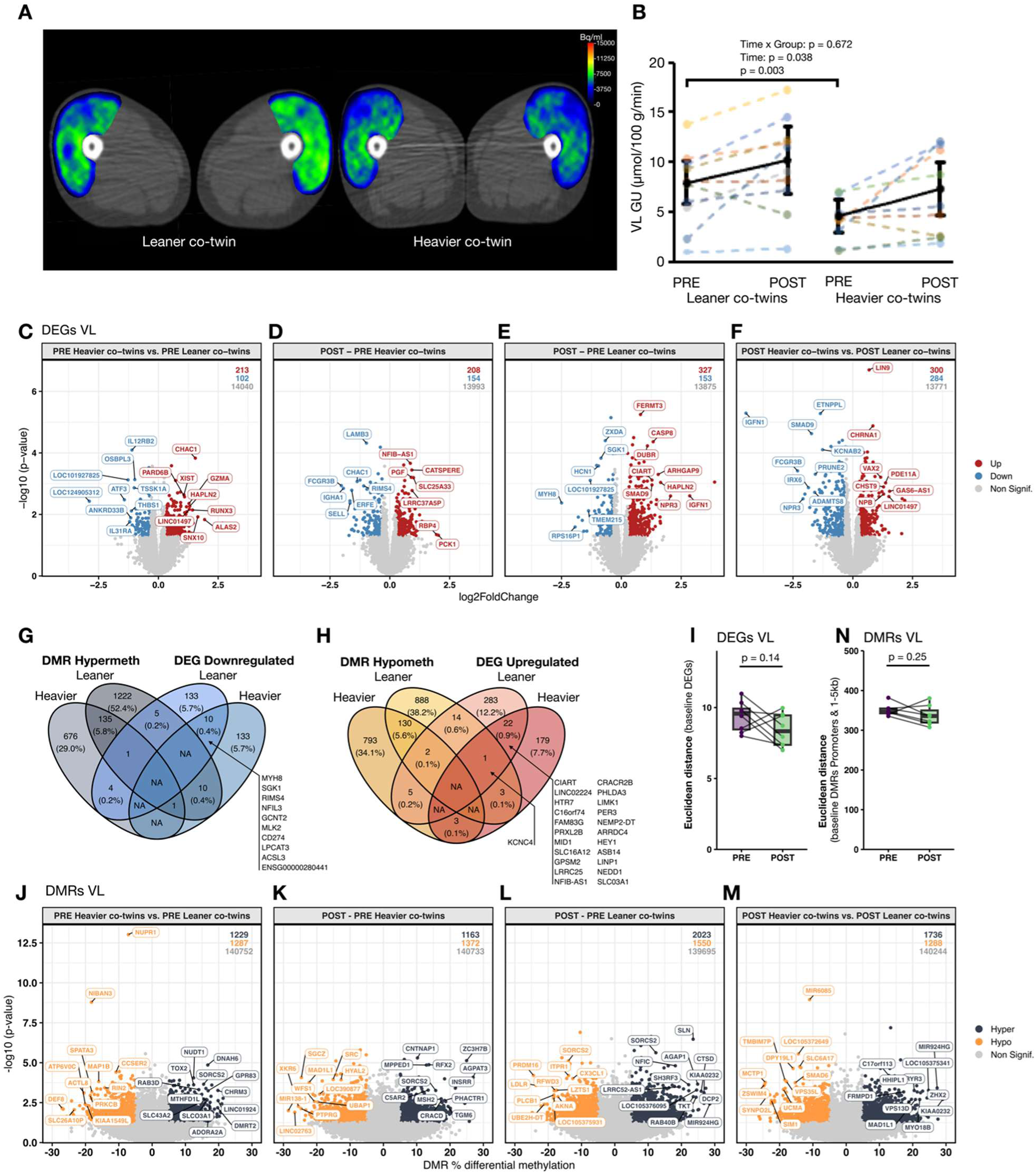
Insulin-stimulated glucose uptake (GU), gene expression and DNA methylation in vastus lateralis (VL) muscle. A) Representative [^18^F]FDG-PET/CT-images of quadriceps femoris in a leaner and heavier co-twin at baseline.s B) Insulin-stimulated glucose uptake in vastus lateralis (VL) muscle in leaner and heavier co-twins at PRE and POST intervention. Bolded line depicts model based mean with 95% confidence intervals. Co-twins share the same colour in the figure. C) Volcano plot of differentially expressed genes (DEGs) between heavier and leaner co-twins before exercise training intervention (PRE). D) Volcano plot of DEGs in response to exercise training intervention in heavier and E) leaner co-twins. F) Volcano plot of DEGs between heavier and leaner co-twins after exercise training intervention (POST). In C-F, the cut-off for statistical significance was set at nominal p ≤ 0.05 with ≥20% difference between co-twins (B&E) or ≥20% change following exercise training (D&E). Top DEGs ranked by XIAO score are annotated in the volcano plots. Displayed values indicate the number of upregulated, downregulated, and non-significant genes per comparison. G) Venn diagram of overlap between downregulated DEGs and differentially hypermethylated regions (DMRs) at gene expression regulatory regions (promoters and regions 1–5 kb upstream of transcription start sites). H) Venn diagram of overlap between upregulated DEGs and hypomethylated DMRs at gene expression regulatory regions following exercise training. I) Euclidean distance in principal component space depicted as boxplots (median, IQR) showing within twin pair divergence at baseline (PRE) and after exercise training (POST); statistical significance was tested by paired t-test. J) Volcano plot of DMRs between heavier and leaner co-twins before exercise training intervention (PRE). K) Volcano plot of DMRs in response to exercise training intervention in heavier and L) leaner co-twins. M) Volcano plot of DMRs at gene expression regulatory regions between heavier and leaner co-twins after exercise training intervention (POST). In J-M the cut-off for the statistical significance was set at nominal p ≤ 0.05 with either ≥5% difference between heavier and leaner co-twins (J&M) or ≥5% change following exercise training (K&L). Top DMRs ranked by XIAO score are annotated in the volcano plots. Displayed values indicate the number of hypermethylated, hypomethylated, and non-significant regions in each comparison. N) Euclidean distance in principal component space based on baseline DMRs in promoters and 1-5kb from transcriptional start site, depicted as boxplots (median, IQR) showing within twin pair divergence at baseline (PRE) and after exercise training (POST); statistical significance was tested by paired t-test.

### 3.10 Distinct and Extensive Exercise-Induced DNA Methylation Changes in Skeletal Muscle

To evaluate whether transcriptional reprogramming in VL coincided with epigenetic changes, we next investigated differential DNA methylation between co-twins. At baseline, 2,516 differentially methylated regions (DMRs) were identified in VL skeletal muscle between heavier and leaner co-twins, including 1,229 hypermethylated and 1,287 hypomethylated regions (Figure 5J). Following exercise training, leaner co-twins exhibited a greater degree of epigenetic remodeling (3,573 DMRs) compared with heavier co-twins (2,535 DMRs), primarily driven by hypermethylation (2,023 regions in leaner vs. 1,163 in heavier twins; Figure 5K & L). Of the exercise-induced DMRs, 6.2% (137 regions) were hypermethylated and 6.4% (132 regions) were hypomethylated in both groups (Supplementary File 2). Despite, 3,024 DMRs persisting between co-twins post-training (Figure 5M), only 98 overlapped with baseline differences (Supplementary Figure 4A) (25 hypomethylated and 33 hypermethylated, Supplementary Figure 4B), indicating that exercise training induced largely distinct DNA methylation responses in each group. At gene regulatory regions (promoters and 1-5 kB upstream of TSSs), 701 baseline DMRs were detected (336 hypermethylated and 365 hypomethylated in heavier co-twins; Supplementary Figure 3C). Post training, heavier co-twins demonstrated 335 hypermethylated and 360 hypomethylated regions (Supplementary Figure 3D), while leaner co-twins exhibited 657 hypermethylated and 497 hypomethylated regions (Supplementary Figure 3E). Only 59 training-induced regulatory-region DMRs overlapped between groups (29 hypermethylated, 30 hypomethylated; Supplementary Figure 3E & F). Interestingly, exercise training did not lead to a reduced epigenetic divergence between co-twins in VL (Figure 5N), in contrast to what we observed in ASAT (Figure 2N).

### 3.11 Limited Epigenetic Overlap but Distinct TF Activity Changes in Skeletal Muscle

To explore regulatory mechanisms underlying gene expression changes in VL skeletal muscle, we examined genes showing both differential methylation and expression (DmDEGs). At baseline, 11 DmDEGs were identified across all annotated regions, regulated by 14 DMRs (Figure 6A). These included two hypermethylated-downregulated genes (*SLCO3A1* and *DYNC1L1*) and three hypomethylated-upregulated genes (*KIAA1549*, *RAC2* and *RNF125*) in heavier versus leaner co-twins (Figure 6A). For training induced responses, heavier co-twins exhibited 11 DmDEGs, including hypomethylated-upregulated *HR*, *ZNF22* and *AGBL1* (Figure 6B). Leaner co-twins demonstrated 43 DmDEGs enriched for transcriptional regulation, protein glycosylation and neuronal signalling, including *TAL1*, *WWOX*, *HDAC9*, *GMDS* and *KCNC4* (Figure 6C). Post training, 49 DmDEGs were detected, regulated by 55 DMRs (7 hypomethylated-upregulated; 16 hypermethylated-downregulated; Figure 6D), with no overlap between baseline and post training DmDEGs. These findings suggest that while DMRs were abundant, only a few percent overlapped with DEGs. Full DmDEG lists and genomic region mapping are provided in Supplementary Files 3.1 and 3.2.

**Figure 6.**
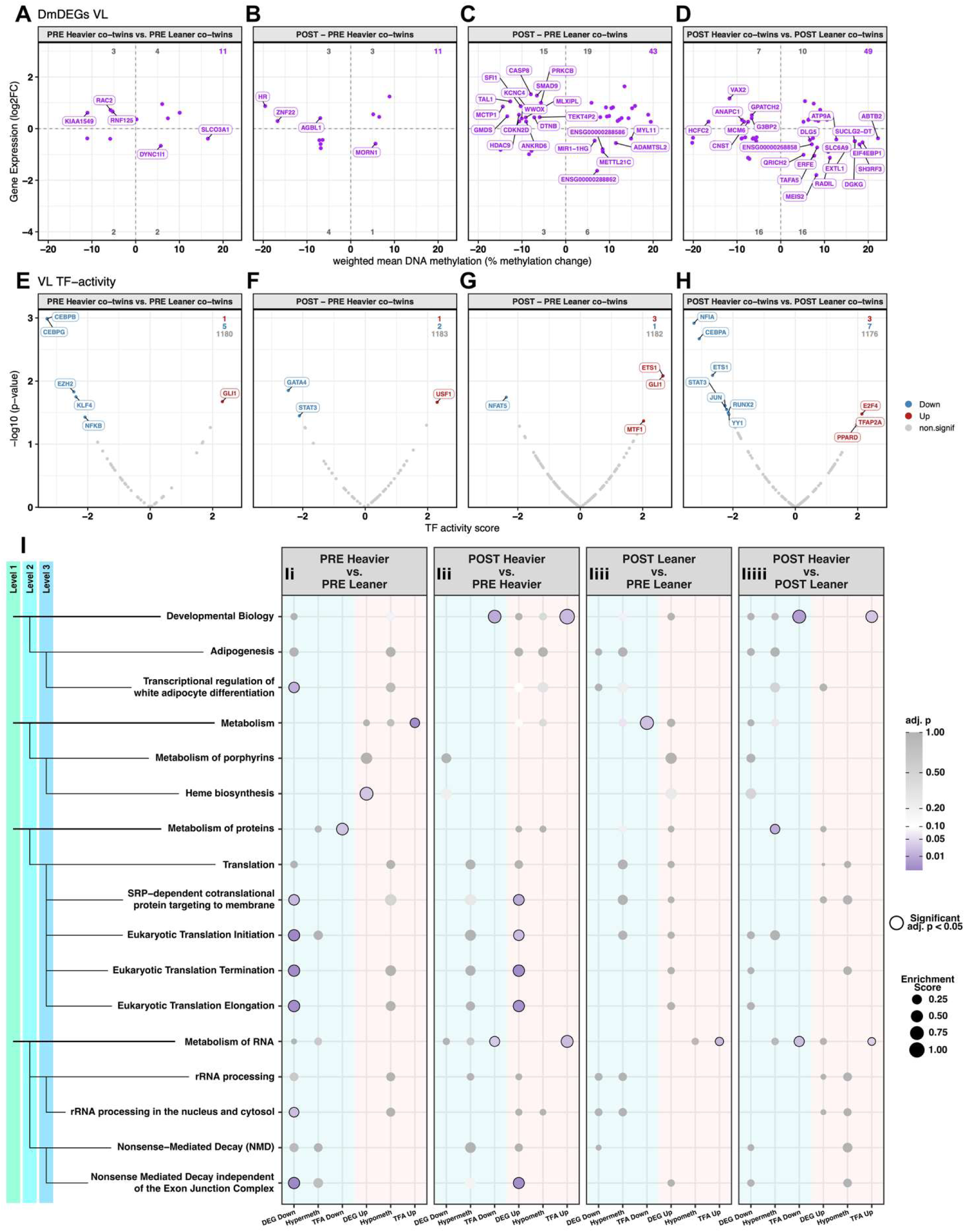
Identification of differentially methylated and expressed genes (DmDEGs), transcription factor activity (TFA), and Reactome pathway enrichment analysis in vastus lateralis (VL) muscle. A) Volcano plot of differentially methylated and expressed genes (DmDEGs) between heavier and leaner co-twins before exercise training intervention (PRE). B) Volcano plot of DmDEGs in response to exercise training intervention in heavier and C) leaner co-twins. D) Volcano plot of DmDEGs between heavier and leaner co-twins after exercise training intervention (POST). In A-D, for genes with more than one significant DMR in a given contrast, we assigned each DMR a weight proportional to the absolute methylation difference, the negative base-10 logarithm of the nominal p-value, and an inverse distance-to-transcription-start-site term, and calculated the weighted mean methylation difference. E) U-plot depicting the difference in transcription factor activity ((TFA) estimated from gene expression data using decouplR)) between heavier and leaner co-twins before intervention (PRE). F) U-plot depicting the difference in TFA in response to exercise training in heavier and G) leaner co-twins. H) U-plot depicting the difference in TFA between heavier and leaner co-twins after the exercise intervention (POST). Ii) A bubble plot showing the top up– and downregulated Reactome pathways identified by differential gene expression between heavier and leaner co-twins before exercise training intervention (PRE). For the top enriched terms identified by gene expression, the role of DNA methylation and TFA were also assessed. Iii) A bubble plot showing the effect of exercise training on the gene expression, DNA methylation and TFA for the top enriched terms identified in (Ii) in heavier and Iiii) leaner co-twins. Iiv) A bubbleplot showing the difference in gene expression, DNA methylation and TFA between heavier and leaner co-twins identified at (Ii) after the exercise training intervention (POST). The colour and the size of the each bubble depicts the degree of the statistical significance and enrichment score, respectively. Bubbles circled with black solid line depict statistically significant (adj. p ≤ 0.05) terms in each comparison.

To further investigate regulatory drivers, transcription factor (TF) activity was inferred from RNA-seq data. VL muscle showed modest TF activity differences at baseline, with increased activity of one TF (*GLI1*) and decreased activity of five TFs (*CEBPB*, *CEBPG*, *EZH2*, *KLF4* and *NFKB*) in heavier co-twins (Figure 6E). Post training within groups, heavier co-twins demonstrated decreased activity of *GATA4* and *STAT3*, and increased activity of *USF1* (Figure 6F), while leaner co-twins exhibited reduced *NFAT5* activity and increased activity of *ETS1, GLI1* and *MTF1* (Figure 6G). Post-training, heavier co-twins exhibited three TFs (*E2F4*, *TFAP2A* and *PPARD*) with increased and seven TFs (*NFIA*, *CEBPA*, *ETS1*, *JUN*, *STAT3*, *RUNX2* and *YY1*) with decreased activity compared with leaner co-twins (Figure 6H). Overall, TF activity changes in VL were modest and spread across a heterogeneous set of factors, including TFs linked to inflammatory and stress-related signaling (for example, *STAT3*) and lipid/oxidative metabolism (for example, *PPARD*). Complete TF-activity lists can be found in Supplementary File 4.

### 3.12 Pathway and Gene-Level Adaptations in Skeletal Muscle and Their Association to Insulin Sensitivity

To explore the biological relevance of DEGs associated with obesity and influenced by exercise training, we performed Reactome pathway enrichment analysis. In VL muscle, heavier co-twins exhibited baseline upregulation of pathways related to heme biosynthesis, whereas pathways involved in RNA processing and protein translation were downregulated (Figure 6Ii). These differences were not explained by DNA methylation or TF activity on pathway level (Figure 6Ii). We next investigated whether exercise training modified the Reactome terms that differed in VL at baseline between heavier and leaner co-twins. Following exercise training, heavier co-twins demonstrated increased expression of genes related to RNA processing and protein translation, although these changes were not accompanied by DNA hypomethylation on pathway level or increased transcription factor activity (Figure 6Iii). The top 10 up/downregulated Reactome pathways induced by exercise training, independent of baseline differences between co-twin groups for both DEGs and DMRs in VL of At baseline, expression of RNA processing and protein translation pathways correlated negatively with systemic inflammation (hs-CRP) and positively with whole-body insulin sensitivity (M-value; Figure 7A). Post-training improvements in skeletal muscle insulin sensitivity were associated with increased expression of these pathways when both co-twin groups were analyzed together (Figure 7B). These findings suggest that enhanced RNA transcriptional and protein translational capacity may associate with improved metabolic function, especially in skeletal muscle of heavier co-twins, following exercise training. To further explore these associations, we identified the individual genes whose training-evoked expression changes in VL muscle showed the strongest correlation with improvements in whole body and skeletal muscle insulin sensitivity. For M-value, the top negatively correlated genes included: *PNPLA2*, *HMGCS2*, *NLRC3, ENSG00000261167* and *CHD9NB*, whereas strongest positive correlation were observed for: *JKAMP*, *PNRC1*, *ZSCAN21*, *NR3C1* and *CNOT10* (Figure 7C). For skeletal muscle insulin-stimulated glucose uptake, the top negative correlations were identified for *CCNJ*, *HYKK*, *SERP2*, *MTA3* and *TMEM135,* while *CTU1*, *PMM1*, *RRP1*, *ASAH2B* and *ENSG00000213062* showed the strongest positive correlations (Figure 7D). These gene-level associations highlight potential molecular drivers of exercise training-induced improvements in skeletal muscle insulin sensitivity and warrant further study.

**Figure 7.**
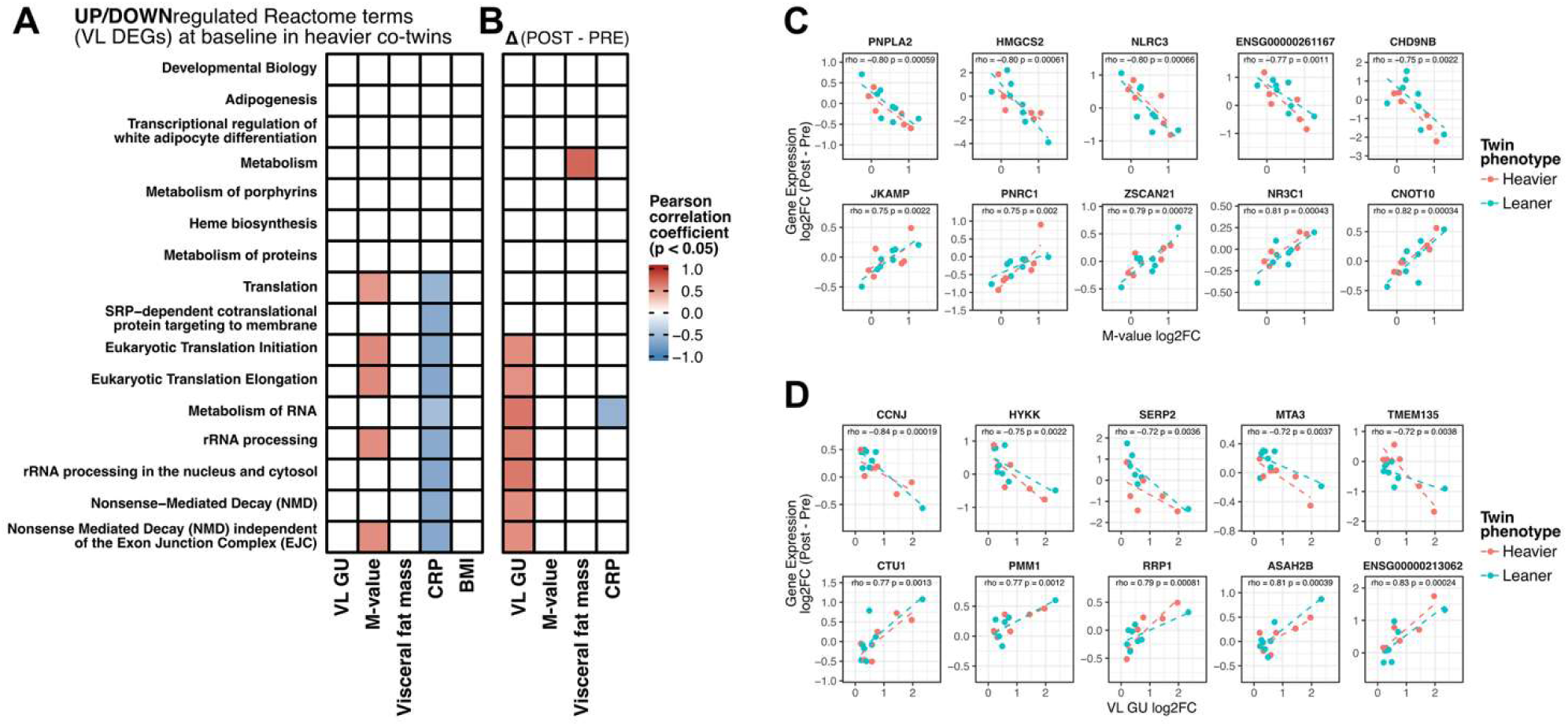
Correlations between gene expression in vastus lateralis (VL) muscle and key phenotype measures. A) A correlation heatmap showing Pearson correlations between Reactome terms (up– or downregulated at gene expression level in heavier compared with leaner co-twins before intervention) and key phenotype outcome measures (insulin-stimulated glucose uptake (GU) in vastus lateralis (VL), whole-body insulin sensitivity (M-value), visceral fat mass, BMI and high-sensitive C reactive protein (CRP)). B) Correlation heatmap showing Pearson correlations between the exercise training intervention induced change (“Δ post – pre”) in Reactome term enrichment at gene expression level and the change (Δ post – pre) in key phenotype outcome measures. Scatterplots depicting the top 5 positively and top 5 negatively correlated (Spearman) individual genes with C) M-value and D) VL GU. In A-D) Only significant correlations (nominal p ≤ 0.05) are colored in the heatmap. The colour intensity depicts the correlation coefficient value. Co-twin groups were pooled for the Δ post – pre correlation analyzes (A-D).

## 4 DISCUSSION

This study in MZ twin pairs discordant for BMI demonstrates that six months of exercise training significantly improved whole-body insulin sensitivity, primarily through adaptations in skeletal muscle rather than abdominal subcutaneous adipose tissue (ASAT), regardless of weight loss. Despite similar exercise training-induced improvements in systemic and skeletal muscle insulin sensitivity in heavier and leaner co-twins, exercise training elicited distinct molecular responses especially in ASAT, reprogramming the heavier co-twins’ transcriptomic signature.

At baseline, heavier co-twins exhibited increased expression of genes related to inflammation (adaptive and innate immune pathways), cell cycle regulation and collagen degradation by matrix metalloproteinases, consistent with previous reports of obesity-driven adipose tissue remodeling[6,8,43,44]. These pathways correlated negatively with whole-body, skeletal muscle and ASAT insulin sensitivity, and positively with BMI and visceral fat mass. Notably, macrophage-related genes such as *TM4SF19*, *TREM2*, and *SPP1*, expressed by lipid-associated macrophages (LAMs), were among the most upregulated, aligning with evidence that LAMs expand during adipose tissue growth[45,46]. Exercise training markedly attenuated these inflammatory and proliferative transcriptional signatures in heavier co-twins, accompanied by downregulation of LAM-related genes (e.g., *TM4SF19*, *TREM2*, *CHIT1*, *SPP1*, *ITGAX*, *CCL3*) as well as inflammatory Reactome terms[45,46]. These changes were supported by differentially methylated and expressed genes (DmDEGs) that mapped to immune system at pathway level as well as decreased transcription factor activity, indicating that epigenetic and transcriptional regulatory mechanisms may play a context-specific role in the suppression of inflammatory and proliferative gene expression in heavier co-twins.

Furthermore, at baseline, heavier co-twins demonstrated decreased expression of genes related to mitochondrial respiration, a finding that is consistent with previous studies at the transcriptional[6,8] and protein level[47]. Although one previous study reported favorable effects of exercise training on ASAT mitochondrial function in women with obesity[15], exercise training had no effect on gene expression related to mitochondrial metabolism in the present study, in line with earlier findings showing no effect of 8 weeks of HIIT in middle-aged obese and T2D men[47]. Similar findings of no change or decrease in mitochondrial gene expression have been observed after diet-induced weight loss, an intervention known to improve whole-body insulin sensitivity[48,49]. However, bariatric surgery-induced weight loss seems to improve aerobic metabolism in ASAT, suggesting that the improvement in ASAT mitochondrial metabolism may be dependent on the metabolic milieu for surgery[49]. In this study no reduction in body fat mass was observed, which may explain why no increase in mitochondrial gene expression was observed.

At baseline, compared with leaner co-twins, heavier co-twins exhibited decreased expression of genes related to brown and beige adipocyte differentiation, collagen biosynthesis, protein translation and RNA processing. In heavier co-twins, exercise training upregulated all these pathways except brown and beige adipocyte differentiation showed a trend for upregulation. Brown and beige adipocyte differentiation and RNA processing were positively associated with whole-body insulin-sensitivity, while collagen formation showed a negative association, suggesting their importance in the regulation of insulin-mediated glucose metabolism. Previous studies have shown that exercise training can upregulate markers of adipose tissue browning, yet this warrants more mechanistic research due to reports of contradictory findings[50,51]. Interestingly, compared with leaner co-twins, heavier co-twins exhibited lower expression and intron hypermethylation of *PRDM16*, which is a master regulator of adipocyte browning[52]. Importantly, exercise training increased the expression of *PRDM16* alongside intron region hypomethylation in heavier co-twins suggesting a molecular mechanism for adipose tissue browning by exercise training as also at the pathway level exercise training tended to upregulate adipose beiging/browning. Furthermore, 8 weeks of HIIT was shown to upregulate translation and RNA-splicing pathways at the protein level in middle-aged T2D men[47]. The above results agree with our findings and suggest that exercise training may restore, to some extent, the obesity-dysregulated protein translation pathways. Despite this, the transcriptional improvements induced by exercise training did not translate into increased ASAT glucose uptake, consistent with previous studies[20,31,33]. This indicates that favorable adipose tissue remodeling by exercise training may contribute to systemic metabolic health indirectly, through reduced inflammation rather than through enhanced adipose tissue insulin-stimulated glucose uptake.

At the gene level, approximately 5% of the obesity-associated DEGs overlapped with directional methylation changes across all annotated regions (hypermethylated-downregulated and hypomethylated-upregulated); intron region methylation showing the highest degree of overlap with gene expression. The hypomethylated-upregulated genes mapped among other pathways to immune system and cell cycle pathways, indicating that a subset of these differentially expressed genes are modulated by context-specific DNA methylation. Additionally, the hypermethylated-downregulated genes mapped among other pathways to lipid metabolism and small molecule transport suggesting they were regulated by DNA methylation.

Compared with ASAT, VL exhibited less extensive transcriptomic differences at baseline and following training, especially in heavier co-twins. Heavier co-twins demonstrated upregulation of heme biosynthesis pathways, while RNA processing and protein translation pathways were downregulated. These differences seemed to be independent of DNA methylation or TF activity at the pathway level, indicating a significant role of another regulatory mechanism such as chromatin accessibility and/or histone modification in driving adiposity related molecular dysregulation. Exercise training reversed this pattern, increasing expression of RNA processing and protein translation-related genes alongside elevated TF activity, suggesting that enhanced transcriptional and protein synthetic capacity could be a potential mechanism for improved insulin sensitivity observed in skeletal muscle. Baseline expression of RNA processing and protein translation pathways also correlated negatively with systemic inflammation (hs-CRP) and positively with whole-body insulin sensitivity, while post-training improvements in skeletal muscle insulin sensitivity were associated with increased expression of these pathways.

Gene-level correlations identified in VL that, in response to exercise training, the change in *JKAMP*, *PNRC1*, *ZSCAN21*, *NR3C1* and *CNOT10* expression were the most positively associated with M-value improvements, whereas *PNPLA2*, *HMGCS2*, *NLRC3* showed the highest negative associations, implicating that genes belonging to lipid metabolism, stress response and myogenesis pathways were associated with improvements in whole-body insulin sensitivity. Interestingly, inflammatory genes such as *FCGR3B*, *IGHA1*, *SELL*, *FPR1*, *S100A8*, *VNN2* and *S100A9* were among the most downregulated in heavier co-twins post-training, suggesting anti-inflammatory effects in skeletal muscle despite modest overall transcriptomic changes. Additionally, *ALAS2*, a key gene in heme biosynthesis, was highly upregulated at baseline in heavier co-twins, consistent with obesity-related iron dysregulation[53–56]. Exercise tended to normalize *ALAS2* expression that could potentially improve iron homeostasis and therefore muscle metabolic health. These genes therefore warrant further investigation in the context of skeletal muscle insulin sensitivity.

In terms of regulatory drivers underlying these improvements, both tissues exhibited extensive DNA methylation differences at baseline and in response to training. However, only a small proportion of DEGs showed concordant directional change in DNA methylation and gene expression, accounting for 5.6% and 2.2% of the baseline DEGs in ASAT and VL, respectively. Among the overlapping DmDEGs, intronic DMRs showed the highest overlap, especially in ASAT, indicating that epigenetic regulation should be further studied beyond cis-regulatory regions. Additionally, the training induced changes in both co-twin groups showed similar degree of overlap (heavier co-twins ASAT: 6.5% and VL: 1.9%; leaner co-twins ASAT: 3.2% and VL: 5.6%) for directional methylation and gene expression. Taken together, these findings suggest that, although DNA methylation remodeling was widespread, it accounted for only a limited fraction of the transcriptional differences associated with obesity and exercise training, indicating that other regulatory mechanisms likely play a prominent role in the observed gene expression responses.

Several factors may have introduced variability into our findings and therefore limitations should be acknowledged: 1) Although the mean BMI difference between co-twins was substantial (7.6 kg/m²) and the heavier co-twins’ average BMI (36.7 kg/m2) was within class II obesity, intra-pair differences ranged from 2.2 to 18.4 kg/m², and the leaner co-twins had BMIs, on average, within the overweight range, potentially attenuating between-pair contrasts. 2) Diet was not strictly controlled; food diary records suggested no statistically significant differences in energy intake at baseline between leaner and heavier co-twins or from PRE to POST, but underreporting, especially in individuals with obesity, is well documented[57]. 3) Biopsies were obtained in a post-prandial state without standardized meal, potentially adding noise to transcriptome and methylome profiles. 4) Sample size was smaller than originally planned due to recruitment challenges and biopsy refusals, potentially limiting power to detect subtle molecular changes, particularly in skeletal muscle. Finally, our analyzes focused on DNA methylation as the primary epigenetic layer, other regulatory layers such as histone modifications, chromatin accessibility, and non-coding RNAs were not assessed. Protein-level validation and single-cell omics were beyond the scope of the current study; thus, the observed transcriptomic changes require further confirmation at the protein and cellular level in future work although our results agree closely with a previous study examining the effect of obesity/T2D and exercise training on the scWAT proteome[47].

Future work should address these limitations by expanding sample size and ensuring greater BMI contrast between co-twins, implementing strict dietary and biopsy timing controls to reduce confounding variables, and incorporating single-cell transcriptomics and methylomics to resolve cell-type-specific adaptations in adipose and skeletal muscle tissue. Multi-omics approaches should be extended to include metabolomics, (phospho)proteomics, and chromatin accessibility for a more comprehensive regulatory map. Investigating acute exercise responses would also help to determine whether chronic methylation changes to obesity or prior training, prime transcriptional responsiveness to future exercise. Functional studies are needed to test causal roles of candidate genes and pathways to clarify systemic metabolic crosstalk between tissues and identify potential therapeutic targets.

In conclusion, in monozygotic twin pairs discordant for BMI, six months of combined exercise training improved skeletal muscle insulin sensitivity, while adipose tissue underwent extensive transcriptomic and epigenetic remodeling without changes in insulin-stimulated glucose uptake. In adipose tissue, exercise training reversed obesity-associated inflammatory and proliferative signatures, accompanied by transcription factor activity normalization and context-specific DNA methylation changes. In skeletal muscle, exercise training reversed the obesity-associated RNA processing and protein translation pathways and these terms associated with higher insulin sensitivity, with gene-level correlates nominating potential molecular drivers. These findings underscore tissue-specific mechanisms of exercise adaptation and highlight transcriptional, translational and inflammatory pathways as promising targets for interventions aimed at improving metabolic health. Overall, we demonstrate that exercise training not only improves skeletal muscle insulin sensitivity but also reprograms the adipose tissue transcriptome in young adults with obesity, highlighting tissue-specific molecular adaptations that may underpin the metabolic benefits of regular exercise training in obesity.

## Data statement

Raw and pre-processed RNA-seq and bisulfite sequencing data are available upon reasonable request following peer-reviewed publication. Processed DEG and DMR data are provided in Supplementary Files 1–5. Any other processed data are available upon reasonable request to the corresponding authors for authors that have institutional review board/ethics approval and an institutionally approved study plan.

## CRediT authorship contribution statement

**Jaakko Hentilä:** Conceptualization, Data curation, Formal analysis, Investigation, Methodology, Visualization, Writing – original draft, Project administration

**Max Ullrich:** Data curation, Formal analysis, Investigation, Methodology, Software, Validation, Visualization, Writing – original draft

**Ronja Ojala:** Investigation

**Thibaux Van der Stede**: Resources, Software

**Marja Heiskanen:** Investigation

**Sanna Honkala:** Investigation

**Martin S. Lietzén:** Investigation

**Mika Helmiö:** Supervision, Investigation

**Olli Eskola:** Resources

**Johan Rajander:** Resources

**Eliisa Löyttyniemi:** Methodology

**Riikka Lautamäki:** Investigation

**Heidi Virtanen:** Investigation

**Kalle Koskensalo:** Investigation

**Olli J. Heinonen:** Investigation, Resources

**Kirsi Pietiläinen:** Resources

**Jaakko Kaprio:** Resources

**Riikka Kivelä:** Conceptualization, Methodology, Formal analysis, Resources, Supervision, Validation, Writing – review and editing

**Adam P. Sharples:** Conceptualization, Resources, Formal analysis, Methodology, Supervision, Validation, Writing – review and editing

**Jarna C. Hannukainen:** Conceptualization, Project administration, Funding acquisition, Methodology, Supervision, Validation, Writing – review and editing

## Data Availability

All data produced in the present study are available upon reasonable request to the authors

## ACKNOWLEDGEMENTS

This study was supported by Finnish Functional Genomics Centre, University of Turku and Åbo Akademi and Biocenter Finland”. The authors want to thank all the volunteers who participated in this study, the personnel of Turku PET Centre and Paavo Nurmi Centre for their excellent assistance. The authors also thank Sini Hentilä (MSc.) for analyzing the food diaries.

