## Supplementary figures for "Exercise Training Improves Skeletal Muscle Insulin Sensitivity and Reprograms the Adipose Transcriptome in Heavier Monozygotic Twins": Supplementary figures.pdf

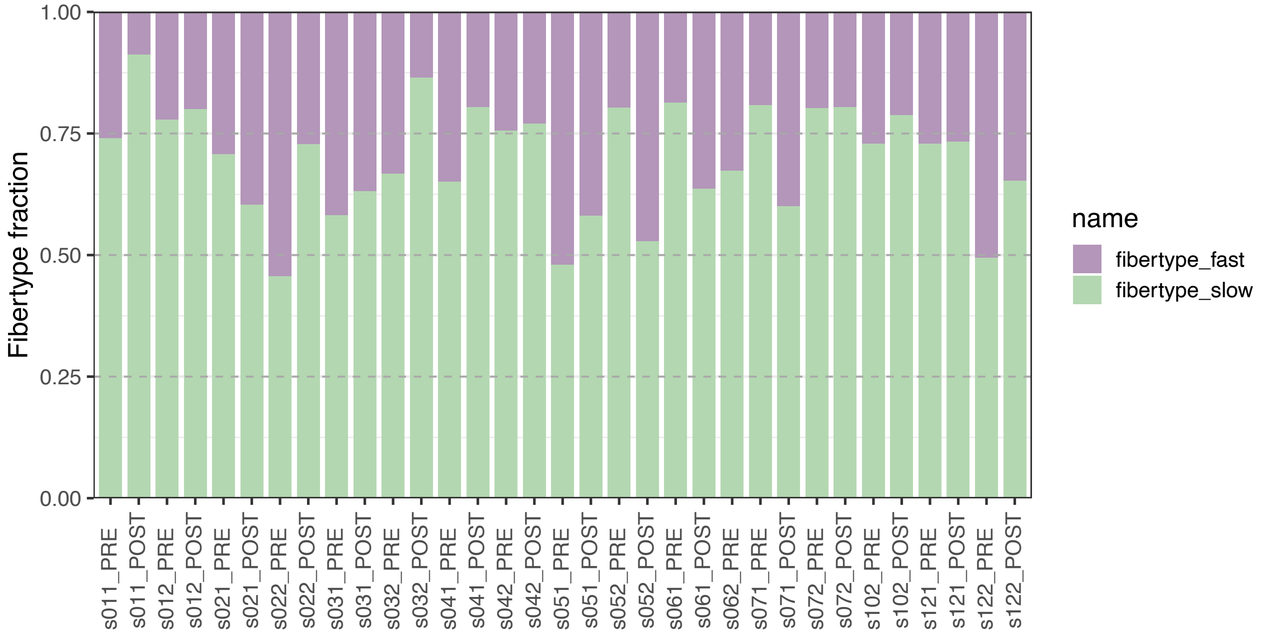


**Supplemental Figure 1.** Deconvoluted Muscle Fiber Type proportions Slow/Fast based on FibeRtypeR applied to skeletal muscle RNAseq data.

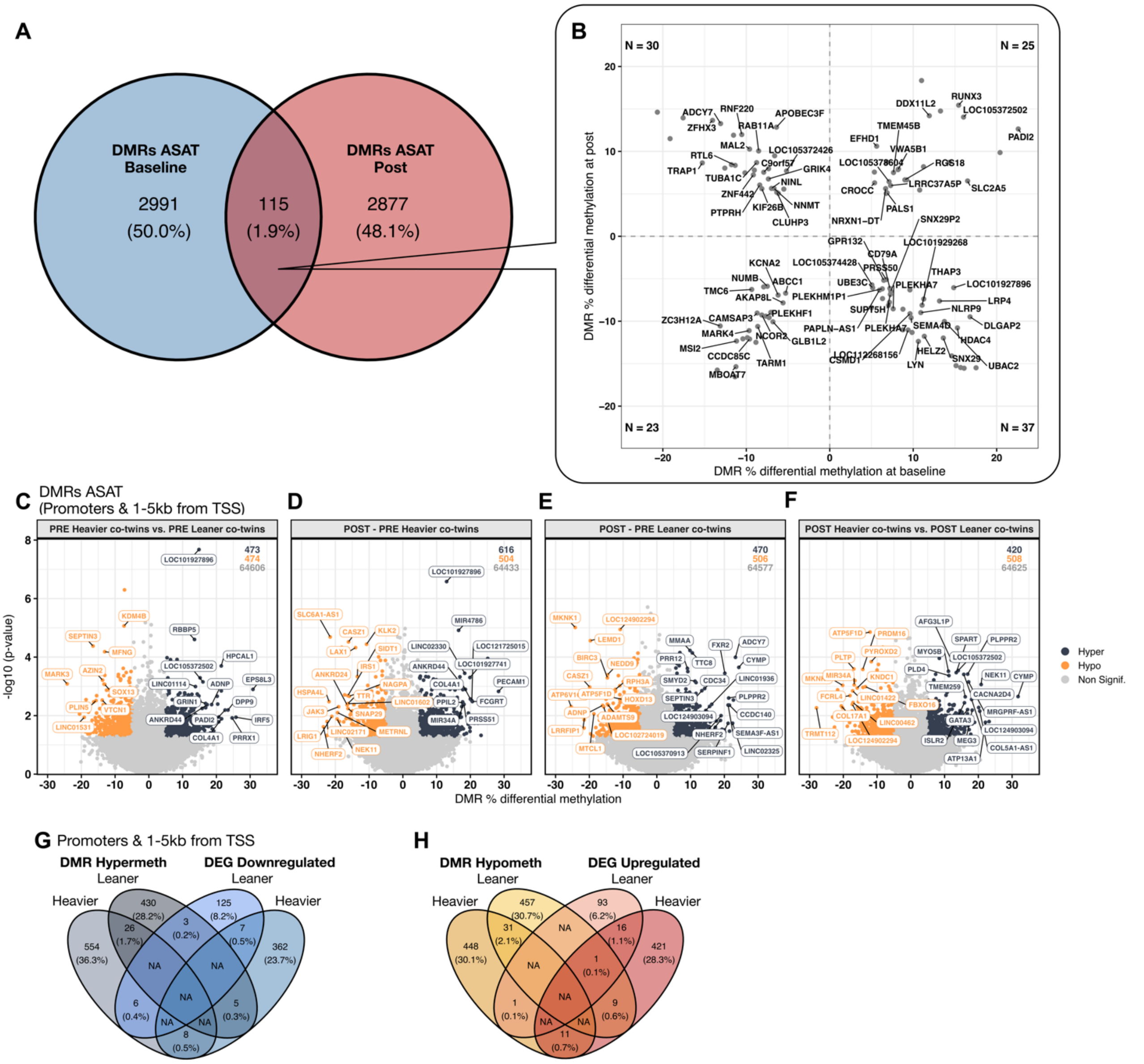


**Supplemental Figure 2. Overlapping Baseline and Post DMRs, and Promoter and 1-5kb from transcriptional start site DNA methylation in abdominal subcutaneous adipose tissue (ASAT).** (A) Venn diagram of overlapped DMRs between heavier and leaner co-twins at baseline and at post in ASAT, overlapped by exact region coordinates. (B) Dot plot of the overlapping DMRs from A, labelled by annotated gene name. Unlabelled points are intergenic DMRs. (C-F) Volcano plots of DMRs in gene regulatory regions (promoters and 1-5kb from TSS) between C) heavier and leaner co-wins at baseline, Post - Pre in D) heavier and E) leaner co-twins, and between F) heavier and leaner co-twins after the exercise training intervention. G-H) Venn diagrams of overlapping directional DMRs in promoters and 1-5kb from TSS and DEGs in ASAT between heavier and leaner co-twins, either (G) hypermethylated -downregulated or (H) hypomethylated-upregulated.

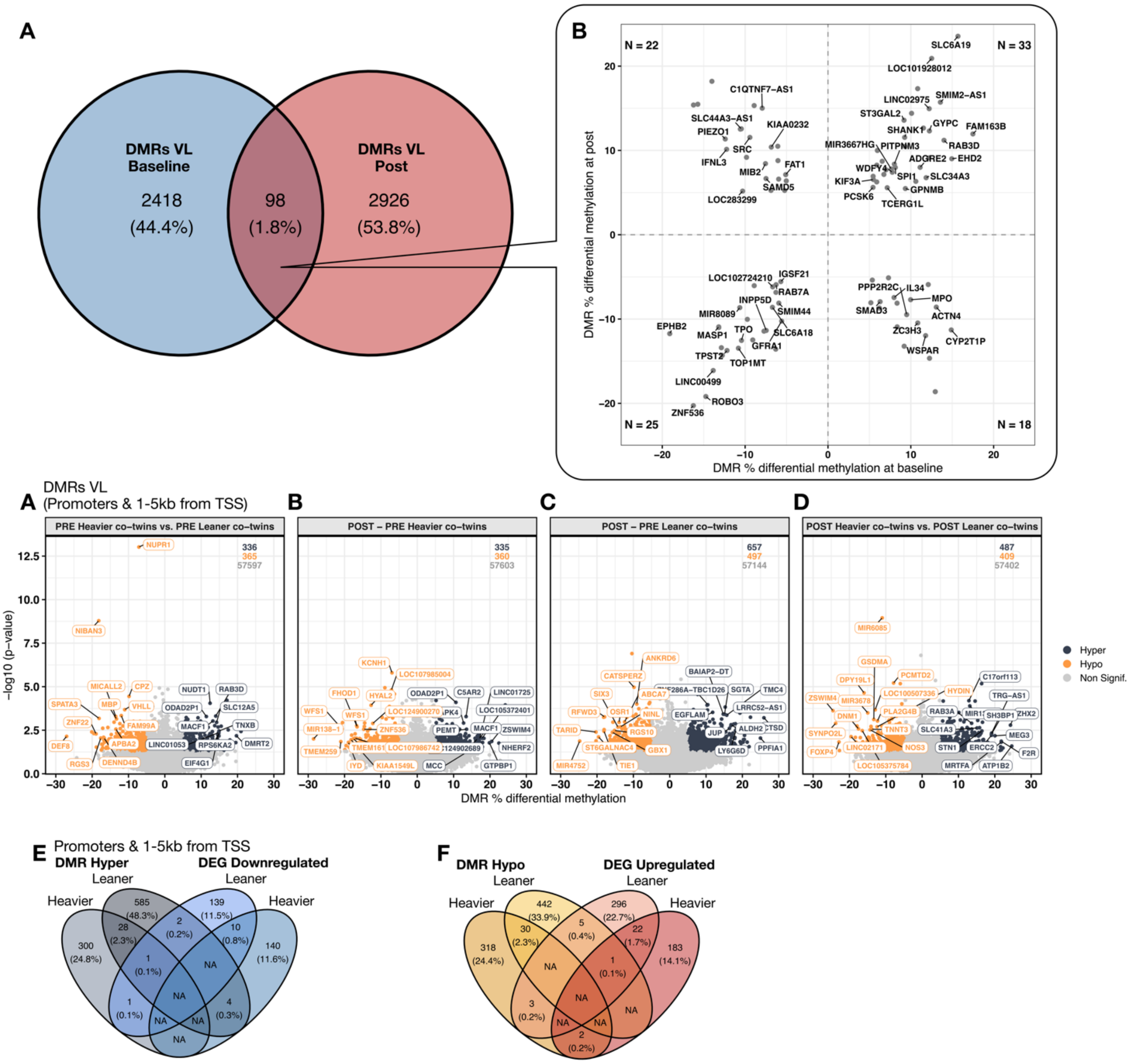


**Supplemental Figure 3. Overlapping Baseline and Post DMRs, Promoter and 1-5kb from transcriptional start site DNA methylation in vastus lateralis (VL).** (A) Venn diagram of overlapped DMRs between heavier and leaner co-twins at baseline and at post in VL, overlapped by exact region coordinates. (B) Dot plot of the overlapping DMRs from A, labelled by annotated gene name. Unlabelled points are intergenic DMRs. (C-F) Volcano plots of DMRs in gene regulatory regions (promoters and 1-5kb from TSS) between C) heavier and leaner co-twins at baseline, Post - Pre in D) heavier and E) leaner co-twins, and between F) heavier and leaner co-twins after the exercise training intervention. G-H) Venn diagrams of overlapping directional DMRs in promoters and 1-5kb from TSS and DEGs in VL between heavier and leaner co-twins, either (G) hypermethylated -downregulated or (H) hypomethylated-upregulated.

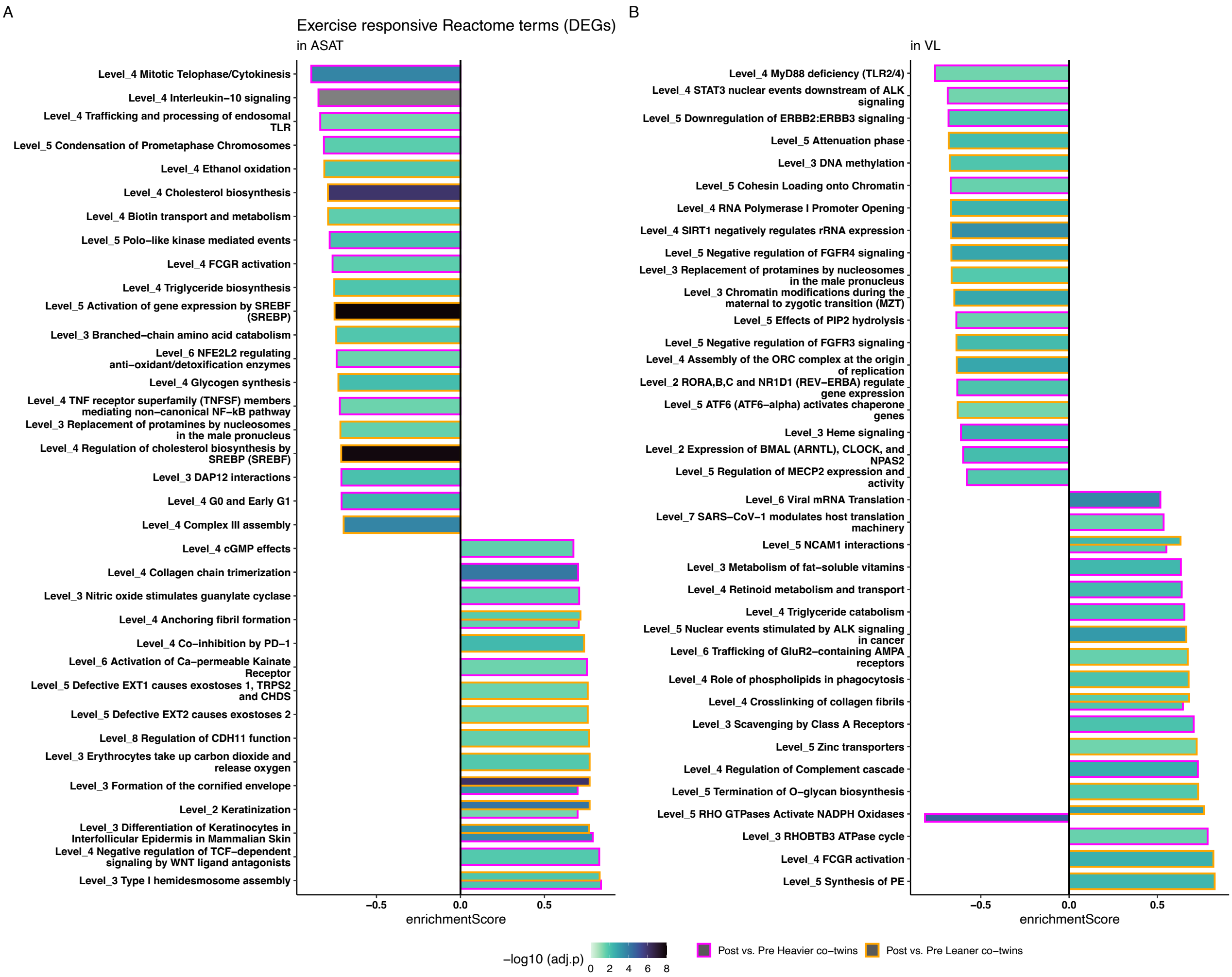


**Supplemental Figure 4. Reactome pathway enrichment analysis of exercise-responsive (Post -Pre) genes (DEGs) in abdominal subcutaneous adipose tissue (ASAT) and vastus lateralis (VL) muscle.** Top 10 up and down regulated Reactome terms in Leaner and Heavier co-twins significantly associated with exercise response (Post – Pre) are shown for ASAT (A) and VL (B). Bars represent enrichment scores for Post vs. Pre comparisons in heavier (magenta) and leaner (orange) co-twins. Fill intensity corresponds to statistical significance ($-\log_{10}$ BH adjusted p-value), with darker shades indicating stronger significance.

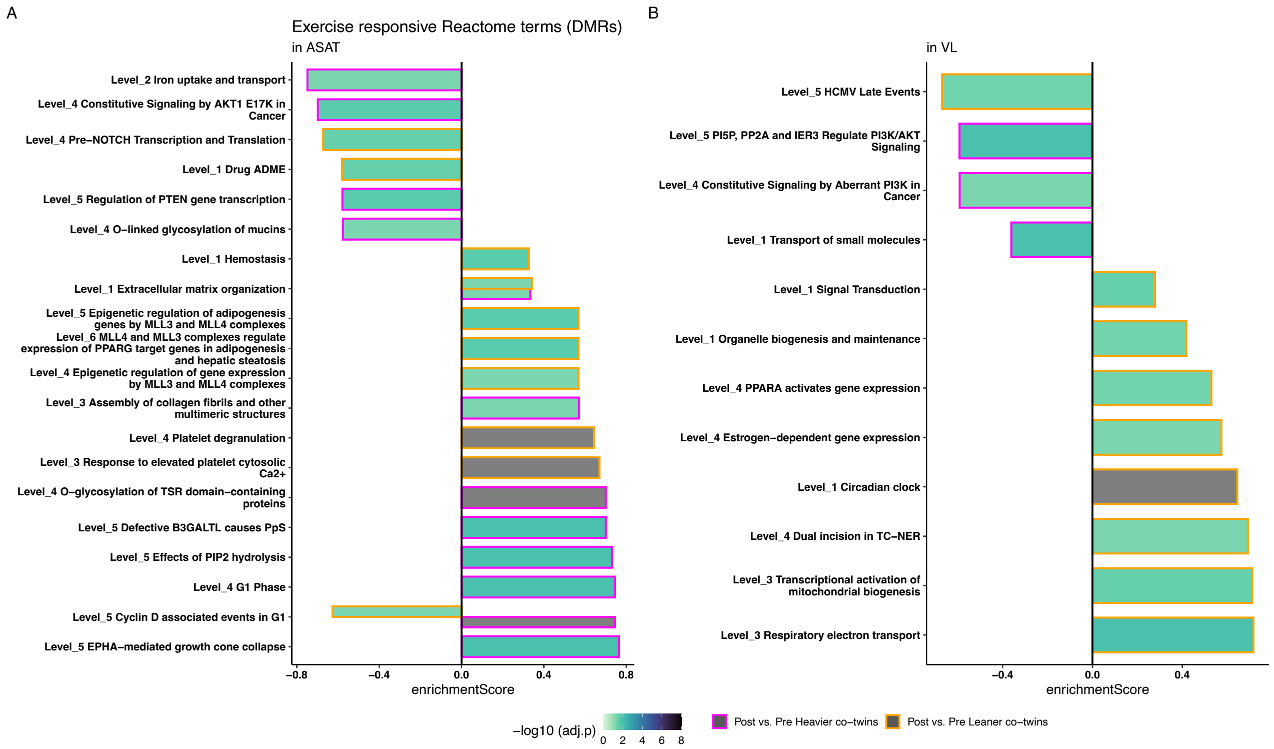


**Supplemental Figure 5. Reactome pathway enrichment analysis of exercise-responsive (Post -Pre) differentially methylated regions (DMRs) in abdominal subcutaneous adipose tissue (ASAT) and vastus lateralis (VL) muscle.** All significantly regulated Reactome terms in Leaner and Heavier co-twins associated with exercise response (Post – Pre) are shown for ASAT (A) and VL (B). Bars represent enrichment scores for Post vs. Pre comparisons in heavier (magenta) and leaner (orange) co-twins. Fill intensity corresponds to statistical significance (−$\log_{10}$ BH adjusted p-value), with darker shades indicating stronger significance.

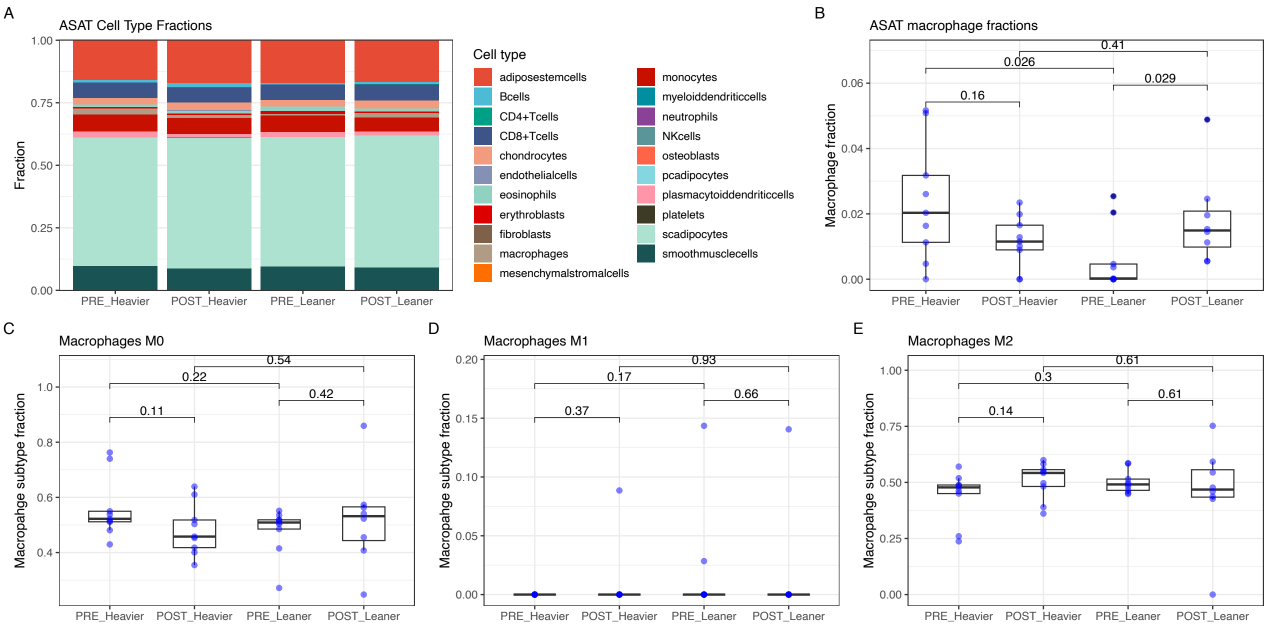


**Supplemental Figure 6**. Cell population deconvolution of ASAT based on gene expression profiles using Cibersort and the adipose tissue specific signature matrix (AT22) and the macrophage profiles from PBMC (LM22). A) All deconvoluted celltype fractions in ASAT. B) Deconvoluted ASAT macrophage fractions in ASAT. C-E) Deconvoluted Macrophage subtypes M0, M1 & M2 based on gene expression profiles in the LM22 reference dataset.
